# Genetic correlates of vitamin D-binding protein and 25 hydroxyvitamin D in neonatal dried blood spots

**DOI:** 10.1101/2022.06.08.22276164

**Authors:** Clara Albiñana, Zhihong Zhu, Nis Borbye-Lorenzen, Sanne Grundvad Boelt, Arieh S. Cohen, Kristin Skogstrand, Naomi R. Wray, Joana A. Revez, Florian Privé, Liselotte V. Petersen, Cynthia M. Bulik, Oleguer Plana-Ripoll, Katherine L. Musliner, Esben Agerbo, Anders D. Børglum, David M. Hougaard, Merete Nordentoft, Thomas Werge, Preben Bo Mortensen, Bjarni J. Vilhjálmsson, John J. McGrath

## Abstract

The vitamin D binding protein (DBP), encoded by the group-specific component (*GC*) gene, is a much-studied component of the vitamin D system. In a genome-wide association study of DBP concentration in 65,589 neonates, we identified 26 independent loci, 17 of which were in or close to the *GC* gene, with fine-mapping identifying 2 loci on chromosomes 12 and 17 (missense variants within *SH2B3* and *GSDMA,* respectively). When adjusted for key *GC* haplotypes, we found 15 independent loci distributed over 10 chromosomes. Mendelian randomization analyses found evidence consistent with a unidirectional, causal effect of higher DBP concentration and (a) higher 25 hydroxyvitamin D (25OHD) concentration, and (b) a reduced risk of multiple sclerosis and rheumatoid arthritis. A phenome-wide association study in an external dataset confirmed that higher DBP concentration was associated with higher 25OHD concentration and a reduced risk of vitamin D deficiency. Our study provides new insights into the influence of DBP on vitamin D status and a range of health outcomes.

## INTRODUCTION

The vitamin D-binding protein (DBP) is a highly polymorphic protein best known for its role related to the transport of 25 hydroxyvitamin D (25OHD) and 1, 25 dihydroxvitamin D (1,25OHD)^1^. DBP, which is an abundant circulating (plasma) protein structurally related to albumin, is encoded by the group-specific component (*GC*) gene. Haplotypes determined by two missense variants in the *GC* gene (rs7041 and rs4588) determine key isoforms of the DBP protein, which are labelled according to their electrophoretic properties (1s, 1f, 2). Apart from these haplotypes, there are many additional variants in humans^1^.

While DBP also has a range of additional roles (e.g., actin scavenging after tissue injury, C5a- mediated chemotaxis, T-cell response, macrophage activation^2^), most research has focused on the contribution of DBP to overall vitamin D status. It is known from related steroid hormones and their binding proteins, that the concentration of the binding protein can influence the bio-availability of the target hormone. Much of this research has been informed by the free hormone hypothesis^3^, which proposes that the biological activity of a hormone is related to the unbound (i.e., free) rather than the protein-bound concentration in the plasma^1^. With respect to total 25OHD, on average only 0.03% is free, 85% is bound to DBP, and the remainder is bound (less strongly) to albumin^4^. Apart from some tissues which can retrieve protein-bound 25OHD via endocytosis (e.g., distal renal tubules, the placenta), free/unbound 25OHD is thought to be the biologically active fraction. Recently, an individual with a homozygous deletion of *GC* was identified, and shown to have DBP and 25OHD concentrations below the limit of detection (the concentration of 1,25OHD was low but detetable).^5^ Interestingly, the affected individual did not display adverse bone outcomes traditionally associated with vitamin D deficiency. Combined with evidence from transgenic animal models of *GC* knock-outs^6^, this indicates that DBP is not necessary for the transport of 25OHD throughout the body, nor is DBP necessary for general bone health. It is now appreciated that the concentration of DBP can influence the half-life of 25OHD^5, 7^. When the concentration of DBP is lower, then more 25OHD is free/unbound, and this fraction of the total 25OHD is more rapidly transferred to target cells and subsequently catabolized, thus shortening the functional half-life of 25OHD.

Despite concerns about the accuracy of some DBP assays (assays based on monoclonal antibodies have underestimated DBP concentration in African-Americans^8^), it is clear that there is appreciable variation in DBP concentration within groups sorted by DBP isoform type^8^. Some of this variation may be related to genetic factors. To date, we are aware of only one genome-wide association study (GWAS) of DBP concentration, based on 1380 men^9^. This study, which used a monoclonal antibody, identified two genome-wide significant loci, both of which were within the *GC* gene (rs7041, rs705117). Of particular interest, variants in *GC* are also strongly and consistently associated with the concentration of 25OHD^10–13^, indicating a critical role for the binding protein in predicting the total 25OHD concentration. In light of the importance of DBP concentration for influencing vitamin D status, there is a need to undertake GWASs of DBP concentration in larger samples, and to explore analytic methods that can take into account potential isoform-specific assay biases. The summary statistics from this type of study could then be used in Mendelian randomization studies — for example, exploring the relationship between the DBP concentration and 25OHD concentration. In addition, we have a particular interest in exploring the association between DBP and (a) neuropsychiatric disorders^14, 15^, and (b) autoimmune disorders^16–18^.

We measured both DBP and 25OHD concentrations in a large sample of neonatal dried blood spots^19^. Because these samples have previously been genotyped, we were able to undertake a GWAS of DBP. The aims of this study were to (a) describe the distribution and epidemiological correlates of DBP in neonatal dried blood spots, (b) examine single-nucleotide polymorphism (SNP)-based and family-based heritability of DBP, and (c) undertake a GWAS of DBP. Based on the results of the GWAS, we (d) used bioinformatics tools to explore properties of the genome-wide significant loci, (e) used Mendelian randomization to explore the association between DBP and 25OHD concentration, and between DBP and a range of potential vitamin D-related candidate disorders and traits (including neuropsychiatric and autoimmune-related disorders), and (f) conducted a phenome-wide association study (PheWAS)^20^ to examine the relationship between the genetic correlates of DBP and a wide range of health phenotypes.

## METHODS

### Samples

This study was based on the Lundbeck Foundation Initiative for Integrative Psychiatric Research (iPSYCH) sample^19^, a population-based case-cohort design to study the genetic and environmental factors associated with severe mental disorders. The iPSYCH2012 sample is nested within the entire Danish population born between 1981 and 2005 (n=1,472,762). In total, 86,189 individuals were selected; with 57,377 individuals diagnosed with at least one major mental disorder (schizophrenia, bipolar disorder, depression, autism spectrum disorder (ASD), attention deficit hyperactivity disorder (ADHD)) and a random population cohort of 30,000 individuals sampled from the same birth cohort. By design, there were individuals overlapping between the case sub-cohorts and the random population sub-cohort. We also included 4,791 anorexia nervosa cases (AN; ANGI-DK) from the Anorexia Nervosa Genetics Initiative (ANGI)^21^, which has the same design as iPSYCH2012. Henceforth, we refer to iPSYCH2012 as the combined dataset with the ANGI samples. Blood spots for the individuals included in iPSYCH2012 were obtained from the Danish Neonatal Screening Biobank^22^ and subsequently genotyped and assayed for the concentrations of 25OHD and DBP.

### Blood spot extraction

Two 3.2 mm discs of dried blood spots (DBS) were punched into 96 well microtiter plates. Proteins were first extracted in a PBS buffer and stored for further use at -80°C. Subsequently, DNA was extracted according to previously published methods.^23^ After storage the protein extracts were aliquoted and were subjected to DBP and 25OHD analysis. Thus, all experimental data originates from a single DBS extraction. Additional details related to blood spot extraction and storage are provided in Supplementary Methods S1.

### Assay of DBP concentration

The extracts were analyzed with a multiplex immunoassay using U-plex plates (Meso-Scale Diagnostics (MSD), Maryland, US) employing antibodies specific for DBP (HYB249-05 and HYB249-01), as well as measuring complement C3 and C4 (results will be reported in a separate manuscript). The antibodies were purchased from SSI Antibodies (Copenhagen, Denmark). Extracts were analyzed diluted 1:70 in diluent 101 (#R51AD, MSD). Capture antibodies (used at 10 ug/mL as input concentration) were biotinylated in-house using EZ-Link Sulfo-NHS-LC-Biotin (#21327, Thermo Fisher Scientific) and detection antibodies were SULFO-tagged (R91AO, MSD), both at a challenge ratio of 20:1. As a calibrator, we used recombinant human DBP #C953 (Bon Opus, Millburn, NJ, USA). Calibrators were diluted in diluent 101, detection antibodies (used at 1 ug/mL) were diluted in diluent 3 (#R50AP, MSD). Controls were made in-house from part of the calibrator solution in one batch, aliquoted in portions for each plate, and stored at -20°C until use. The samples were prepared on the plates as recommended by the manufacturer, and were read on the QuickPlex SQ 120 (MSD) 4 min after adding 2x Read buffer T (#R92TC, MSD). Analyte concentrations were calculated from the calibrator curves on each plate using 4PL logistic regression using the MSD Workbench software.

Intra-assay variations were calculated from 38 measurements analyzed on the same plate of a pool of extracts made from 304 samples. Inter-assay variations were calculated from controls analyzed in duplicate on each plate during the sample analysis (1022 plates in total). The lower limit of detection was calculated as 2.5 standard deviations from 40 replicate measurements of the zero calibrator. The higher detection limit was defined as the highest calibrator concentration. The lower and upper detection limits for DBP were 2.07 µg/L and 79.8 mg/L respectively, and intra-assay and inter-assay coefficient of variance was 7.6% and 22.4% respectively. Additional details related to pre-analytic variation are provided in Supplementary Methods S2.

### Assay of 25OHD concentration

Detailed methods for the main assay of 25OHD^24^ and an additional method to correct for exposure to bovine serum albumin^25^ have been published elsewhere. We adapted previously published methods^26–28^ in order to assay 25OHD based on protein pellets previously extracted from dried blood spots. In summary, after a liquid-liquid extraction and drying down, the vitamin D metabolites were derivatized with 20 µL of the commercial PTAD reagent. The LC system was a Thermo TLX2 Turboflow system, comprised of a CTC Analytics HTS PAL autosampler, a dual LC system (one Agilent 1200 quartinary and one Agilent 1200 binary pump) and two Thermo Scientific hot pocket column heaters. The LC systems were interfaced with a triple quadrupole mass spectrometer (Thermo Scientific TSQ Quantiva) equipped with a heated electrospray ionization probe.

Intermediate precision was obtained by quantifying the concentration of three stable-isotope-labeled external commercial quality controls with a low, medium and high concentration of each vitamin D metabolite. The percent coefficient of variances ranged from 4.7-7.2%. With respect to the accuracy of the method, we evaluated SRM^®^ 972 sera from the National Institute of Standards and Technology (NIST), finding excellent accuracy (92-105%). The method was able to detect a concentration of both 25OHD2 and 25OHD3 down to approximately 5 nmol/L in the protein extracts. All analyses were based on total 25OHD (the sum of 25OHD2 and 25OHD3). The laboratory participated in the Vitamin D External Assessment Scheme (DEQAS)^29^ during the period of the study. Additional details of the assay and related measures of precision are provided in Supplementary Methods S3.

### Genotyping and quality control

Individuals included in iPSYCH2012 were genotyped using the Infinium PsychChip v1.0 array (Illumina, San Diego, CA, USA). In total, 80,873 individuals were successfully genotyped across 26 waves for ∼550,000 variants^19^. We excluded SNPs with minor allele frequency (MAF) < 0.01, Hardy Weinberg Equilibrium (HWE) *p*-value < 1×10^-5^ or non-SNP alleles (i.e., insertions and deletions, INDELs). 245,328 autosomal SNPs were retained in the backbone set. The backbone set was used to impute the genotypes with the Haplotype Reference Consortium reference panel^30^ following the RICOLPILI pipeline^31^. Imputed best guess genotypes were further filtered for imputation quality (INFO score > 0.8), genotype call probability (P > 0.8), missing variant call rates < 0.05, Hardy–Weinberg equilibrium (HWE) *P*- value ≥ 1×10^-5^ and minor allele frequency (MAF) > 0.01, resulting in 6,091,695 variants remaining.

Darker skin color can reduce actinic production of vitamin D, and because non-European ancestry is associated with variants in DBP (which can influence protein concentration), our primary analyses were in those with European ancestry. We performed principal component analysis (PCA) following Privé *et al*.^32^. The genetic ancestry of the samples was inferred following Privé et al.^33^, where 73,645 individuals were classified as having European ancestry (See Supplementary Methods S4). The genetic relationship matrix (GRM) of the individuals was estimated by *GC*TA v1.93^34^. There were 57,747 unrelated individuals with pairwise coefficient of genetic relationships < 0.05.

### Phenotype distributions and covariates

From the 77,482 individuals with genetic data, 71,944 and 71,212 had DBP and 25OHD measurements respectively. The DBP and 25OHD metabolites were quantified in 1,030 and 1,010 plates, respectively. The quantification plates for DBP and 25OHD explained 11.8% and 55.6% of the phenotypic variance respectively. Note that the sequence of testing followed date of birth, so the marked seasonal variation in 25OHD concentration would be captured in the between-plate variance. We used linear mixed models to pre-regress the effect of the quantification plates from DBP and 25OHD and applied a rank-based inverse-normal transformation (RINT) to the model residuals. The raw distributions of the neonatal DBP and 25OHD can be seen in Supplementary Figure S6. For DBP in the full sample, the mean (and standard deviation) was 2.24 (1.44) µg/L (median and interquartile range: 2.00, 1.19-2.98 µg/L). For DBP in the European subsample, the mean (and standard deviation) was 2.25 (1.44) µg/L (median and interquartile range: 2.01, 1.21-2.99 µg/L). We examined the association between (a) sex, year and month of birth, gestational age, matenal age, and (based on infant genotype) the first 20 principal components (PCs) on (b) 25OHD and DBP concentrations. After, adjusting for plate effect, none of these variables were significantly associated with DBP levels, while month of birth, year of birth, gestational age and maternal age were still significantly associated with 25OHD levels. Additional details for all covariate associations and distributions can be found in the Supplementary Data SD1.

### Genome-wide association study (GWAS) analyses

To identify genetic variants associated with neonatal DBP and 25OHD blood concentrations, we performed a linear mixed model GWAS implemented in fastGWA^35^ on the subset of European ancestry individuals (NDBP = 65,589, N25OHD = 64,988). After pre-adjusting for the quantification plates, we fitted sex, year of birth, genotyping wave and the first 20 PCs as covariates in the model in the DBP genetic analyses, and additionally month of birth, gestational age and maternal age in the 25OHD genetic analyses. In light of the strong influence of the *GC* haplotypes of DBP concentration^9^, and the potential haplotype-related bias in our monoclonal assay^8^, we also performed a GWAS adjusted for the 6 *GC* diplotypes, which were fitted as a covariate in the fastGWA model. Henceforth, we will label the two DBP GWASs and related post-GWAS analyses as (a) DBP (unadjusted GWAS), and (b) DBP_GC (GWAS for DBP adjusted for *GC* haplotypes).

To identify independent associations, we conducted a conditional and joint (COJO; GCTA -- cojo-slct) analysis^36^ using default settings and the European ancestry subset of individuals as LD reference. In addition, we conducted a multi-trait conditional and joint (mtCOJO) analysis^37^ to condition results from the UK Biobank (UKB) 25OHD GWAS^11^ on (a) DBP and (b) DBP_GC with fastGWA.

The iPSYCH case-cohort study is enriched with individuals with psychiatric disorders (i.e., the cases) but also contains a uniform randomly-selected population-based subcohort. To explore if case-enrichment in the sample may have biased the findings from the GWAS, as a planned sensitivity analysis, we ran the GWAS again only within the population-based sub- cohort. Based on the union of the genome-wide significant loci from the entire case-cohort and the subcohort samples, we examined the correlation between the effect sizes (beta values) using Pearson’s correlation coefficients^38^.

### Heritability and SNP-based heritability

Our sample had 23,126 individuals that shared at least one off-diagonal GRM value > 0.05, of which 6,313 had a (off-diagonal) GRM value > 0.2 with at least one other individual in the sample. We estimated the heritability of both 25OHD and DBP using methods described by Zaitlen et al.^39^, within the subset with European ancestry. This method estimates pedigree-based and SNP-based heritability simultaneously in one model using family data and is implemented in GCTA^34^.

Finally, we estimated the SNP-based heritability using LD score regression^40^, SBayesS^41^ and LDpred2-auto^42^ from the GWAS summary statistics. We also estimated the polygenicity (p) parameter with SBayesS and LDpred2-auto. In order to derive these estimates, we used linear regression GWAS summary statistics from unrelated European individuals (NDBP = 48,842, N25OHD = 48,643) and filtered down to the intersection with the HapMap3 set of variants (https://www.sanger.ac.uk/resources/downloads/human/hapmap3.html).

### Fine-mapping and functional annotation

Fine-mapping of the GWAS summary statistic results was performed using a combination of (a) PolyFun^43^ for computing prior causal probabilities based on functional annotations, and (b) SuSiE^44^ which fine-maps the variants and provides posterior inclusion probabilities (PIPs) and credible sets of variants. First, we estimated truncated per-SNP heritabilities for both our GWAS summary statistics (DBP and DBP_GC) using the L2-regularized S-LDSC method described in PolyFun for the set of coding, conserved, regulatory and LD-related annotations described in Gazal et al.^45^ The LD-scores for these annotations were computed using our subset of European-ancestry individuals belonging to the subcohort (N = 24,324). We then used the truncated per-SNP heritabilities as prior causal probabilities in SuSiE for fine-mapping. We only performed fine-mapping on the genome-wide significant loci on the DBP GWAS summary statistics. The credible sets obtained in SuSie were functionally annotated using the Ensembl Variant Effect Predictor (VEP) v85^46^.

### Out-of-sample genetic risk prediction

We identified 1,529 individuals of near-European ancestry (see Supplementary Methods S5) and used this sample as a pseudo-replication sample to examine the out-of-sample prediction accuracy of polygenic risk scores (PRSs). The PRS for 25OHD was computed with SBayesR^47^ and downloaded from the PGS Catalog (ID PGS000882)^48^. The PRSs for the four phenotypes (DBP, 25OHD and these two adjusted for the *GC* haplotypes) were constructed using SBayesS^41^ and LDpred2-auto^42^ from our set of GWAS summary statistics. We used linear regression GWAS summary statistics (with the sample filtered for relatedness) for the PRS methods. For SBayesS, we used the provided UKB HapMap3 shrunk sparse LD matrix as LD reference. For LDpred2, we used the LD blocks based on the subset of HapMap3 variants provided in the paper as LD reference.

We also calculated PRSs using the independent SNP weights estimated by COJO^36^ and clumping threshold (C+T) method with window size 250kb and *r*^2^ < 0.1 (M = 201,402 SNPs)) and *P*-value thresholds (5×10^-8^, 1×10^-6^, 1×10^-4^, 0.001, 0.02, 0.05, 0.1, 0.2, 0.5, 1). The prediction models examined the phenotypic variance explained (r2) after adjusting for sex, age and the first 20 PCs.

### Genetic correlations

The genetic correlation between 25OHD and DBP was estimated in a bivariate GREML analysis (GCTA --reml-bivar) and from GWAS summary statistics with bivariate LD score regression^49^.

### FUMA, GSMR, SMR and PheWas

Functional Mapping and Annotation of Genome-Wide Association Studies (FUMA)^50^ was used to examine gene-based and gene set analyses. We conducted generalised summary-based Mendelian randomization (GSMR)^37^ to explore the causal relationship between (a) DBP and 25OHD blood concentrations, and (b) between DBP concentration and a range of psychiatric and cognitive phenotypes (schizophrenia, major depression, bipolar disorder, ASD, ADHD, Alzheimer’s disease, educational attainment), and with selected autoimmune disorders (multiple sclerosis, amyotrophic lateral sclerosis, type 1 diabetes [T1D], Crohn’s disease, ulcerative colitis, rheumatoid arthritis). All the relevant GWAS summary statistics are publicly available and references for these are provided in the Supplementary Methods S6. As the effect of DBP and 25OHD on these phenotypes may be driven by pleiotropy, the analyses were conducted with and without applying the heterogeneity in dependent instrument (HEIDI) outlier method, which removes loci with strong putative pleiotropic effects^37^. We randomly sampled 10,000 unrelated European individuals from iPSYCH2012 as the LD reference cohort. We used a Bonferroni-corrected threshold of 3.8×10^-3^ (0.05/13) in the GSMR analysis.

We performed summary-data-based MR (SMR) to identify genes with causal/pleiotropic effects on DBP, using the eQTL data from GTEx v8^51^. For this analysis, we used the same LD reference cohort as used in the GSMR analysis. In total, tTere were 195,904 probes from 49 tissues. We accounted for multiple testing by using a Bonferroni-corrected threshold of 2.6×10^-7^ (0.05/195,904).

The PheWAS analysis was conducted in the UKB using; 1) linear model, *yj* = *xj* + *cj* + *ej* for quantitative traits, or 2) logistic model, logit(*yj*) = *xj* + *cj* + *ej* for dichotomous traits, where *yj* represents phenotype in UKB, *xj* represents the polygenic score of DBP or DBP adjusted for *GC* genotypes, and *cj* represents the covariates. There were 1,149 phenotypes included in the PheWAS analysis, 1,027 diseases, 52 anthropometric and brain imaging measures, 70 infectious disease antigens. The diseases were classified by using the International Classification of Diseases, 10^th^ version (ICD-10) code. The quantitative traits were normalised using RINT with mean 0 and variance 1. The polygenic scores were generated using SBayesR^47^ with the reference LD matrix estimated from 1,145,953 HapMap 3 SNPs in the UKB. PRSs were computed for 348,501 individuals of European ancestry. The individuals were genetically unrelated (relationship < 0.05). The covariates included in the model were sex, age and 20 PCs. The significance threshold used was 4.4×10^-5^ (0.05/1,149).

### Ethics and data approvals

The study was approved by the Danish Data Protection Agency, and data access was approved by Statistics Denmark and the Danish Health Data Authority. Approval by the Ethics Committee and written informed consent were not required for register-based projects [Act no. 1338 of 1 September 2020, section 10 on research ethics for administration of health scientific research projects and health data scientific research projects]. All data were de-identified and not recognizable at an individual level.

## RESULTS

### 25OHD and DBP phenotypes

The dataset consisted of 71,944 and 71,212 individuals who had DBP and 25OHD neonatal blood concentrations respectively, with a subset of 65,694 individuals with data on both DBP and 25OHD. The neonatal concentrations of both 25OHD and DBP were right-skewed (Supplementary Figure S6). For the full sample, the Pearson’s correlation coefficient between 25OHD and DBP was 0.19 (*P*-value < 2.2×10^-16^) (Supplementary Figure S7). As expected, 25OHD concentration showed prominent seasonal fluctuation, but there was no seasonal fluctuation in DBP concentrations (Supplementary Figure S8).

Three main haplotypes were inferred from the two well-characterized loci within the *GC* gene (rs7041 and rs4588; Supplementary Table S1 and S2)^1^. The distributions of DBP and 25OHD for each of the 6 possible haplotype combinations (i.e., diplotypes reflecting the contribution of the different haplotypes on each chromosome) for the European ancestry subsample is shown in Figure 2, (the same figure for the entire sample can be found in the Supplementary Figures S9).

**Figure 1.**
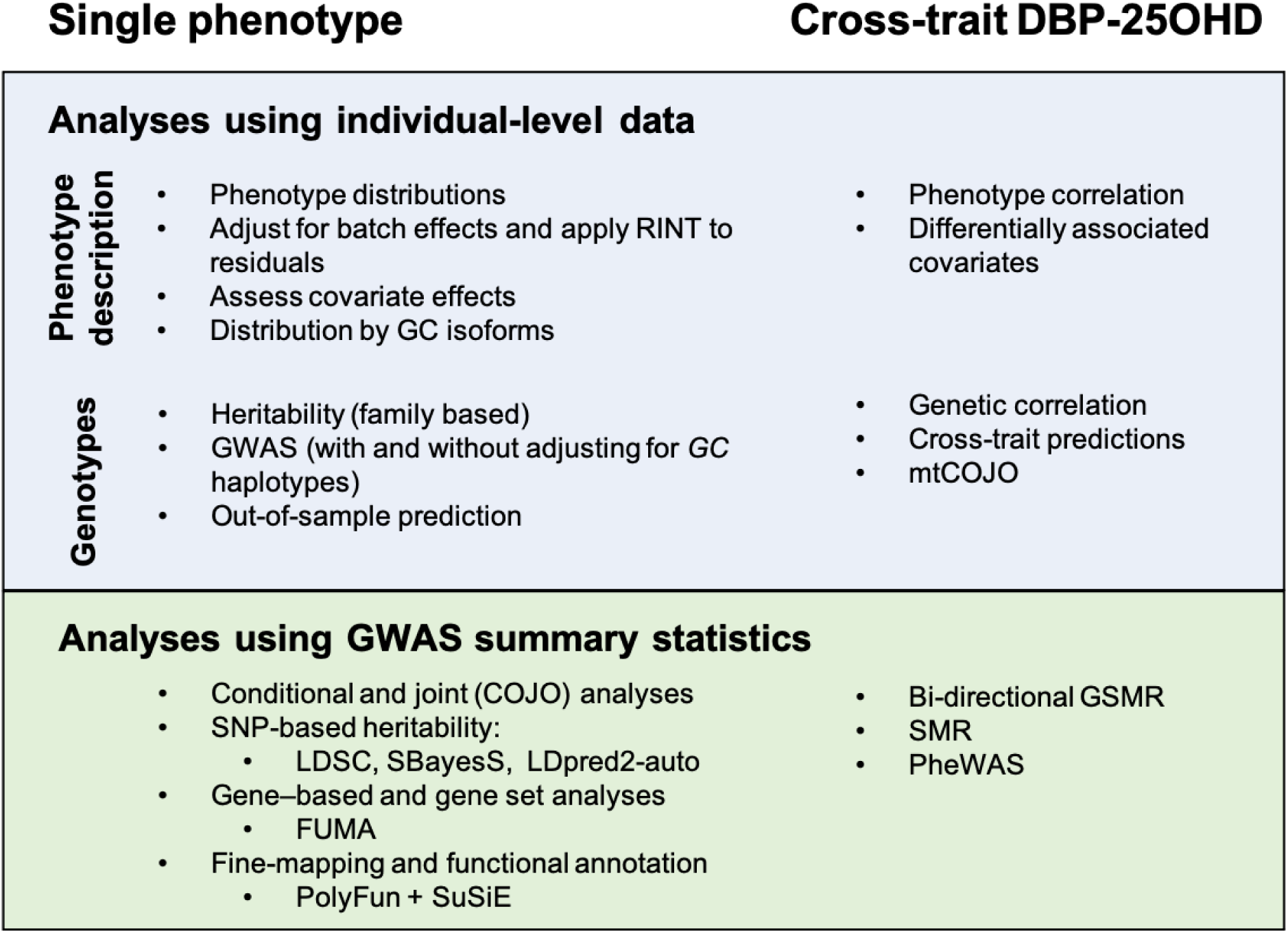
Outline of analyses in this study. Methods figure. FUMA: Functional Mapping and Annotation of Genome-Wide Association Studies, SMR: summary data–based Mendelian randomization, GSMR: Generalised Summary-data-based Mendelian Randomisation, PheWAS: phenome-wide association study, SNP: single nucleotide polymorphism, LDSC: LD-score regression, mtCOJO: multi-trait-based conditional & joint association analysis using GWAS summary statistics. SuSiE: “sum of single effects”.

**Figure 2.**
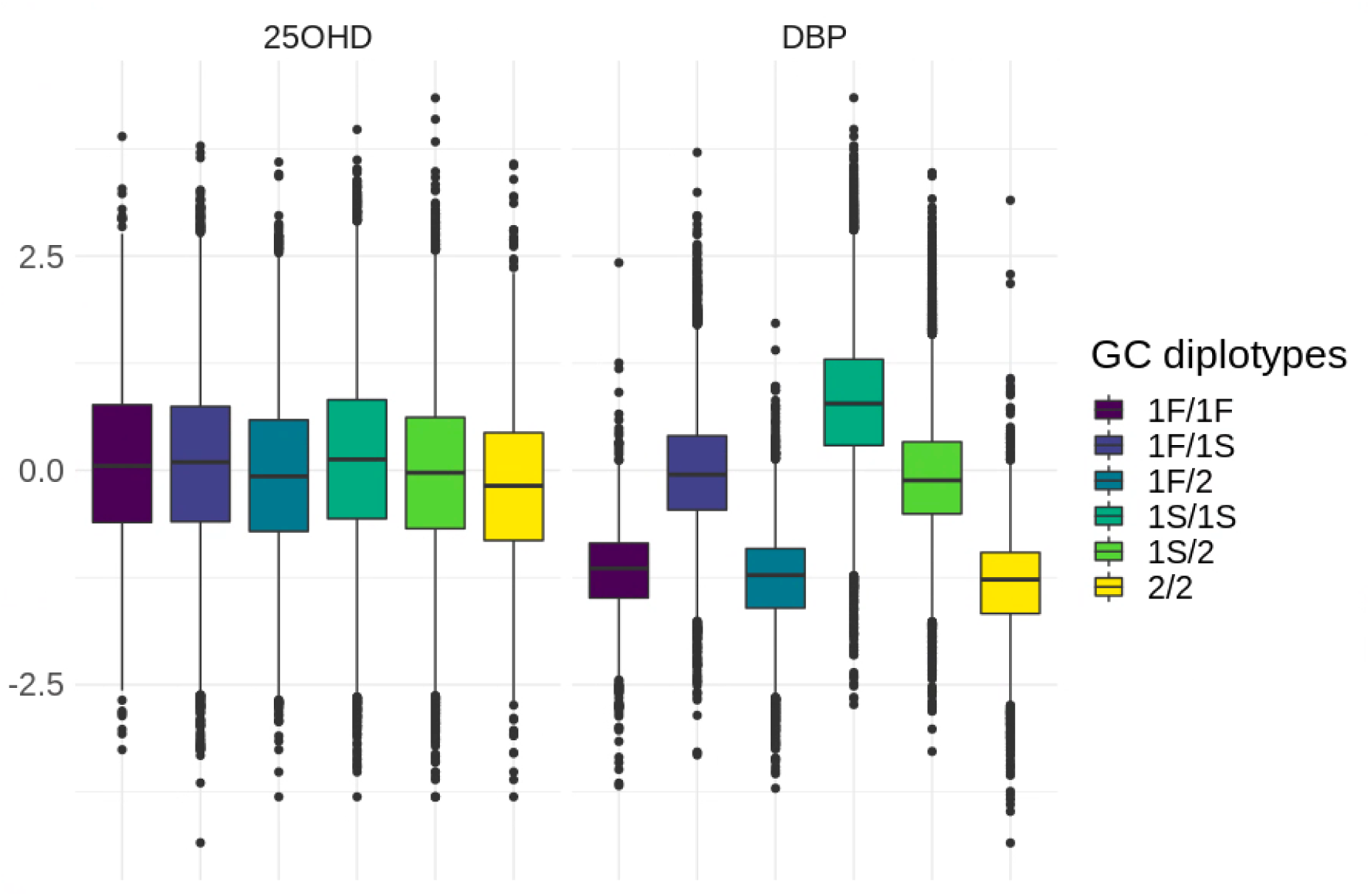
Distributions of transformed 25 hydroxyvitamin D (25OHD) and vitamin D binding protein (DBP) by the six diplotypes, within the European ancestry subgroup. The colour represent the 6 diplotype combinations.

The six *GC* diplotypes were significantly associated with both DBP and 25OHD levels in the full sample and European ancestry subsample (ANOVA *P*-value < 2×10^-16^; Figure 2 and Supplementary Figure S9). In keeping with previous literature based on monoclonal antibodies^8^, the 1S *GC* isoform was associated with higher DBP concentrations. The proportion of DBP variance explained by the *GC* haplotypes was 52.6%, while it was only 0.8% for the 25OHD levels. For completeness, we also show the relative proportion of *GC* haplotypes in the non-European sample. As expected, the 1F isoform was more prevalent in those with African ancestry. (Supplementary Figure S10).

To estimate the heritability of 25OHD and DBP, we used GCTA GREML^39^, and obtained SNP-heritability estimates from GREML, SBayesS and LDpred2. The last two can model sparse genetic architectures, while GREML assumes an infinitesimal architecture. For the family-based heritability estimate we used a set of 6,313 related individuals with coefficient of relationship (r) > 0.2 to at least one other person in the set (all relatives). For 25OHD, the family-based heritability (and standard error) was 0.36 (0.03) and 0.35 (0.03) after adjustment for GC haplotypes (Supplementary Data SD2).

With respect to DBP, the phenotypic variance dramatically declined from 1.0 to 0.47 after the adjustment, because of the substantial contribution of *GC* haplotypes (Figure 3).

**Figure 3.**
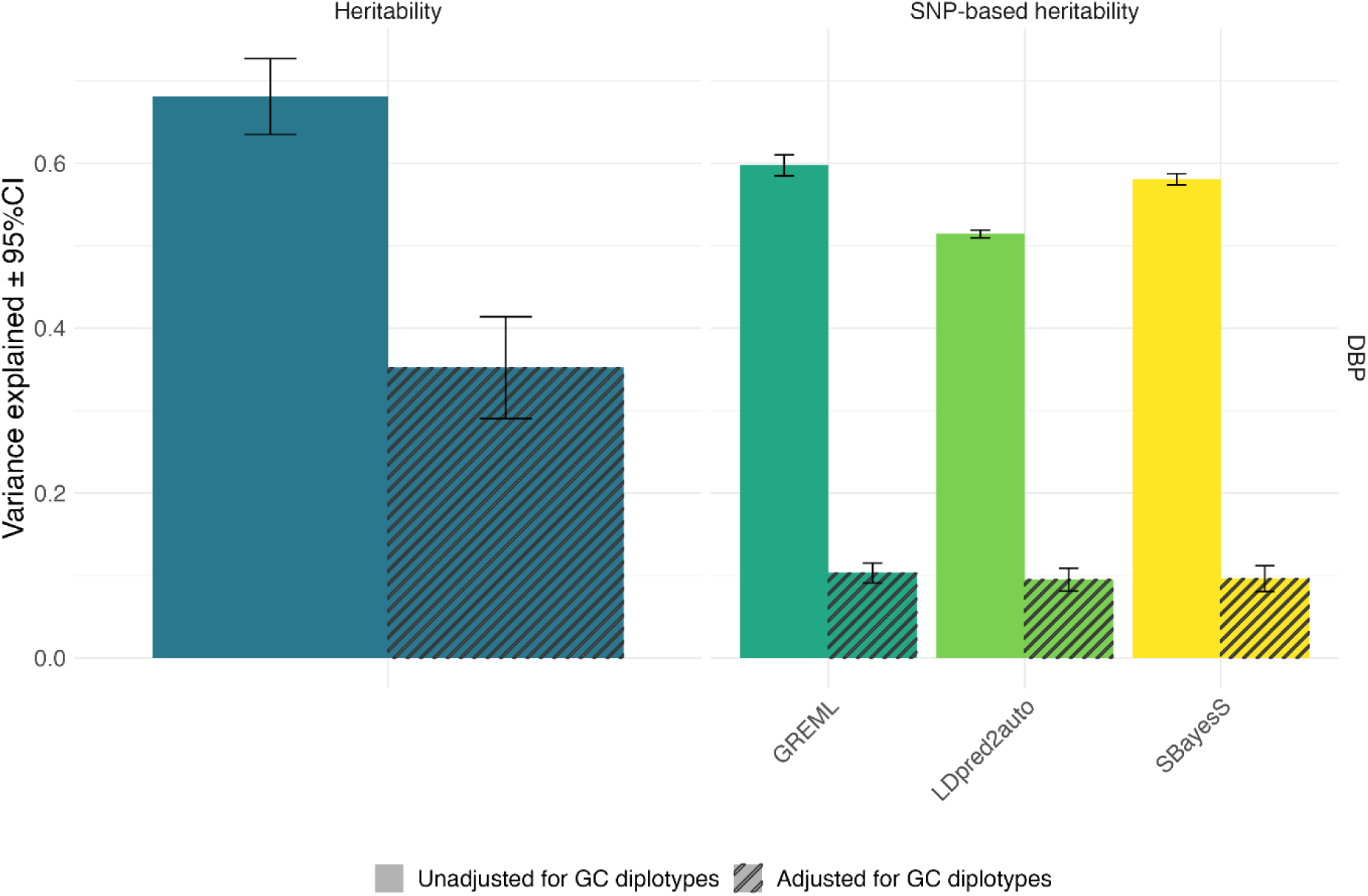
Heritability and SNP-based heritability of DBP. Heritability (left panel) and SNP-based heritability (right panel) estimates) for DBP, with and without adjustment for GC haplotypes (adjusted values shown with cross-hatching). Heritability and SNP-based heritability estimates are presented with 95% confidence interval.

Heritability is a ratio statistic, subject to phenotypic variance. Therefore, we reported the constitutions of heritability. Before the adjustment of GC haplotypes, the heritability (and standard error) of DBP was 0.68 (SE = 0.02), the genetic variance explained by SNPs = 0.58 (SE = 0.01) and the shared environment = 0.10 (SE = 0.02). When adjusted for GC haplotypes, the genetic variance explained by SNPs decreased to 0.05 (SE = 0.002) while the contribution of shared environment was comparable, 0.12 (SE=0.02) (Supplementary Data SD2). These findings indicate that the suggested heritability of DBP is 0.68 and 58% of variance in DBP was captured by SNPs, 53% attributed to *GC* haplotypes and 5% attributed to additional genetic variation. These results are consistent with the estimates from SBayesS and LDpred2-auto (Supplementary Data SD2) and lend weight to the potential informative value of both the DBP and DBP_GC GWASs.

### Genome-wide association study (GWAS) analysis and fine mapping

A total of 6,091,695 SNPs with MAF ≥ 0.01 were tested in the GWAS analysis. Based on GCTA–COJO we identified one independent SNP associated with 25OHD concentration, 26 independent SNPs associated with DBP levels (24 of which were on chromosome 4), and 15 independent SNPs (distributed over 10 chromosomes) associated with DBP levels after adjusting for the *GC* haplotypes (Figure 4). The independent loci for 25OHD located in the *GC* gene on chromosome 4 (rs1352846) had been previously identified^11^. For DBP, we further fine-mapped the genome-wide significant regions in chromosomes 4, 12 and 17 using a combination of PolyFun and SuSiE^43^. For chromosome 4, the key *GC* haplotype- determining rs7041 had a posterior causal probability (PIP) of 1 (Supplementary Data SD11). In the GWAS for DBP_GC, an intergenic locus (rs112704913, chromosome 4: 72,571,221bp, hg19) located 36 kb upstream from the start position of *GC* gene (chromosome 4: 72,607,410-72,669,758bp, hg19, Ensembl) was also identified (PIP = 0.87). From the 26 COJO-independent hits for DBP, 17 loci were in or close to *GC* (9 upstream, 7 within, and 1 downstream of *GC,* all within a 400kb range). For chromosome 12, there was a credible set of 4 SNPs with cumulative PIP > 0.95, where the leading SNP rs3184504 (PIP = 0.5) is a missense variant in *SH2B3*. When adjusting DBP for the GC haplotypes, the fine-mapped results decreased the credible set to 3 SNPs, and the PIP of the missense variant increased to 0.78. This shows how the adjusted GWAS increased the power to fine-map potentially causal variants. For chromosome 17 we observed a similar effect. The fine-mapping algorithm did not output a credible set for this region, and the leading SNP rs56030650 (a missense variant in *GSDMA*) had a low PIP of 0.2. Nevertheless, after adjusting for the *GC* haplotypes, the cumulative PIP of the credible set of 9 SNPs was 0.95, with the missense variant increasing PIP to 0.26. FUMA gene-based analysis also showed that the SNPs in *GC*, *SH2B3* and *GSDMA* were over-represented (Supplementary Data SD3). Additional results are shown in Supplementary Data SD11). Locus zoom plots for these 3 regions are shown in Supplementary Figures SF11, SF12 and SF13.

**Figure 4.**
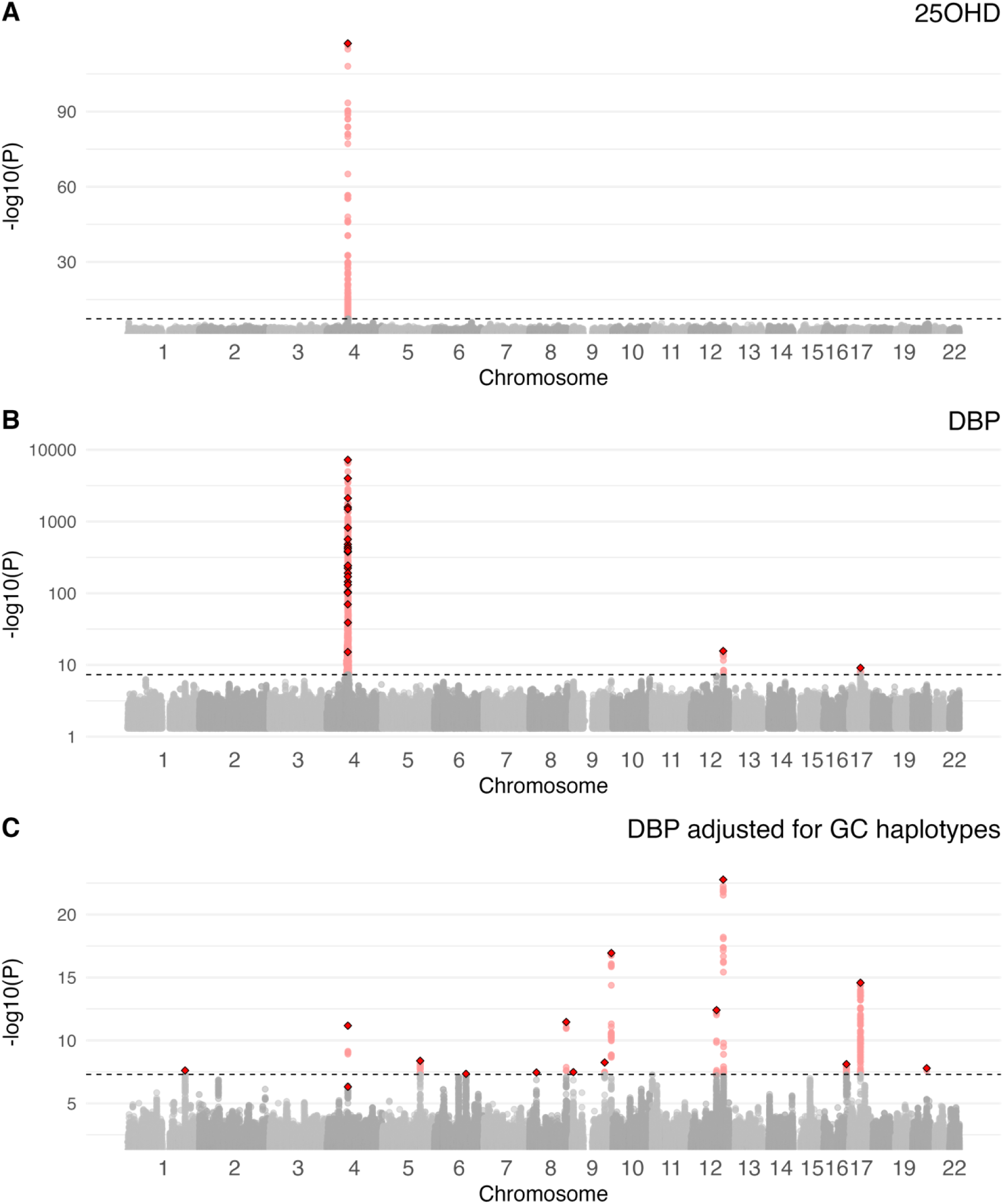
Manhattan plots for 25 hydroxyvitamin D (25OHD), vitamin D binding protein (DBP), and DBP adjusted for GC haplotypes in the iPSYCH case-cohort study. Panel A shows the Manhattan plot for 25OHD, with one prominent peak over the GC gene Chromosome 4. Panel B shows the Manhattan plot for DBP with a very large peak over the GC gene on Chromosome 4. Panel C shows the Manhattan plot for DBP_GC. Note, the Y axis for – log10(p) varies between the panels.

### Replication, out-of-sample genetic risk prediction and sensitivity analysis

We examined if the genetic architecture of 25OHD in our neonatal sample was broadly consistent with that found in the large (n = 417,580) UKB adult sample^11^. Of the 143 genome-wide significant COJO SNPs in the UKB 25OHD GWAS, only the most highly significant one was replicated in our 25OHD sample. However, the Pearson’s correlation coefficient between the allele effect sizes for the union of both GWASs genome-wide significant SNPs (i.e., the significant findings from both UKB and the current sample) was 0.66 (p value < 2.2×10^-16^; Supplementary Figure S11), supporting the hypothesis that the neonatal and adult genetic correlates of 25OHD are broadly comparable.

In order to examine the influence of DBP on 25OHD concentration, we used mtCOJO to condition the UKB 25OHD GWAS summary statistics on our DBP summary statistics. When assessed with and without adjustment for the *GC* haplotypes, we confirmed that the genetic correlates of DBP GWAS were highly influential on 25OHD concentration in an external sample, with only 76 and 79 SNPs out of the 143 COJO SNPs in the UKB 25OHD GWAS remaining genome-wide significant, respectively (Supplementary Data SD4).

With respect to out-of-sample prediction (the European sample predicting into the excluded near-European sample), the proportion of DBP variance explained by the effect of the main SNP rs7048 alone from the DBP GWAS was 54%, while adding more SNPs to the polygenic score (PRS COJO, LDpred2-auto, SBayesS) decreased the r2 to 47%. The maximum variance explained by the P+T PRS was 44% by the smallest threshold p-value < 5e-8, which included 111 SNPs. The proportion of DBP variance explained by the DBP GWAS results adjusted for the GC diplotypes was not significantly different from 0 for any of the PRSs but the SBayesS, which had an r2 of 9% (Supplementary Table S5).

With respect to the planned sensitivity analyses where we compared GWAS findings based on the full case-cohort versus the subcohort only, we found that the SNP effect sizes had a Pearson’s correlation coefficient of 0.99 (*P*-value < 2.2×10^-16^, 3,839 SNPs) for DBP GWAS and of 0.97 (*P*-value < 2.2×10^-16^, 412 SNPs) for the DBP_GC GWAS. This result supports the hypothesis that the findings based on the overall case-cohort sample are comparable to that found in the nested (smaller) general population sample. GWAS findings from the subcohort are shown in Supplementary Data SD6.

### Functional mapping of GWAS findings

We performed gene-based and gene-set analyses, for which results can be found in Supplementary Data SD7 and SD8. As expected, the gene-based analyses identified many genes on chromosome 4 proximal to the *GC* gene. With respect to gene ontology, the top pathway we identified was related to polysaccharide metabolic processes, which may reflect post-translational glycosylation of the DBP protein (a process that influences the properties and elimination of DBP)^4^.

We used SMR to explore the pleiotropic genes that are associated with DBP. Based on both *GC* adjusted and unadjusted summary statistics, loci on chromosome 17 in close proximity to the adjacent genes *MED24* and *GSDMA* were identified (Supplementary Data SD9 and SD10). These results are consistent with the genes identified by the FUMA gene-based analysis.

### GSMR analyses between DBP and 25OHD

The genetic correlation between summary statistics based on GWASs for unadjusted DBP and 25OHD based on bivariate GREML was 58% (SE = 0.05). When estimated at the GWAS summary statistics level with bivariate LDSC regression, it was 34% (SE = 0.09) and 24% (SE = 0.11) when using the unadjusted and GC-adjusted DBP summary statistics respectively, confirming the substantial contribution of DBP to 25OHD concentration (Supplementary Data SD2).

We used bi-directional GSMR to investigate potential causal relationships between DBP and 25OHD. In the following text, we will focus on the findings with HEIDI filtering (which reduces the potential influence of pleiotropy in the analyses; See Supplementary Data SD12 for more details). We found strong support for the hypothesis that high DBP is causal for high 25OHD levels (Figure 5). Concerning the forward GSMR (i.e., DBP predicting UKB 25OHD), we found a highly significant association (*bxy* = 0.08, SE = 0.005, *P*-value = 8.21×10^-55^, *N*SNPs = 40). Concerning the reciprocal (reverse) relationship (UKB 25OHD predicting DBP), there was no significant association (*bxy* = 0.03, SE = 0.02, *P*-value = 0.14, *N*SNPs = 201). After adjusting on the *GC* haplotypes, the general pattern of finding persisted (forward GSMR: *bxy* = 0.06, SE = 0.02, *P*-value = 0.01, *N*SNPs = 10; reverse GSMR: *bxy* = 0.003, SE = 0.01, *P*-value = 0.82, *N*SNPs = 222). Therefore, these results suggest that a 1 SD unit (1.44 ug/L) increase in DBP concentration results in an increase of 0.06-0.08 x SD unit (14.1 nmol/L) of 25OHD concentration (i.e., 0.85-1.13 nmol/L).

**Figure 5:**
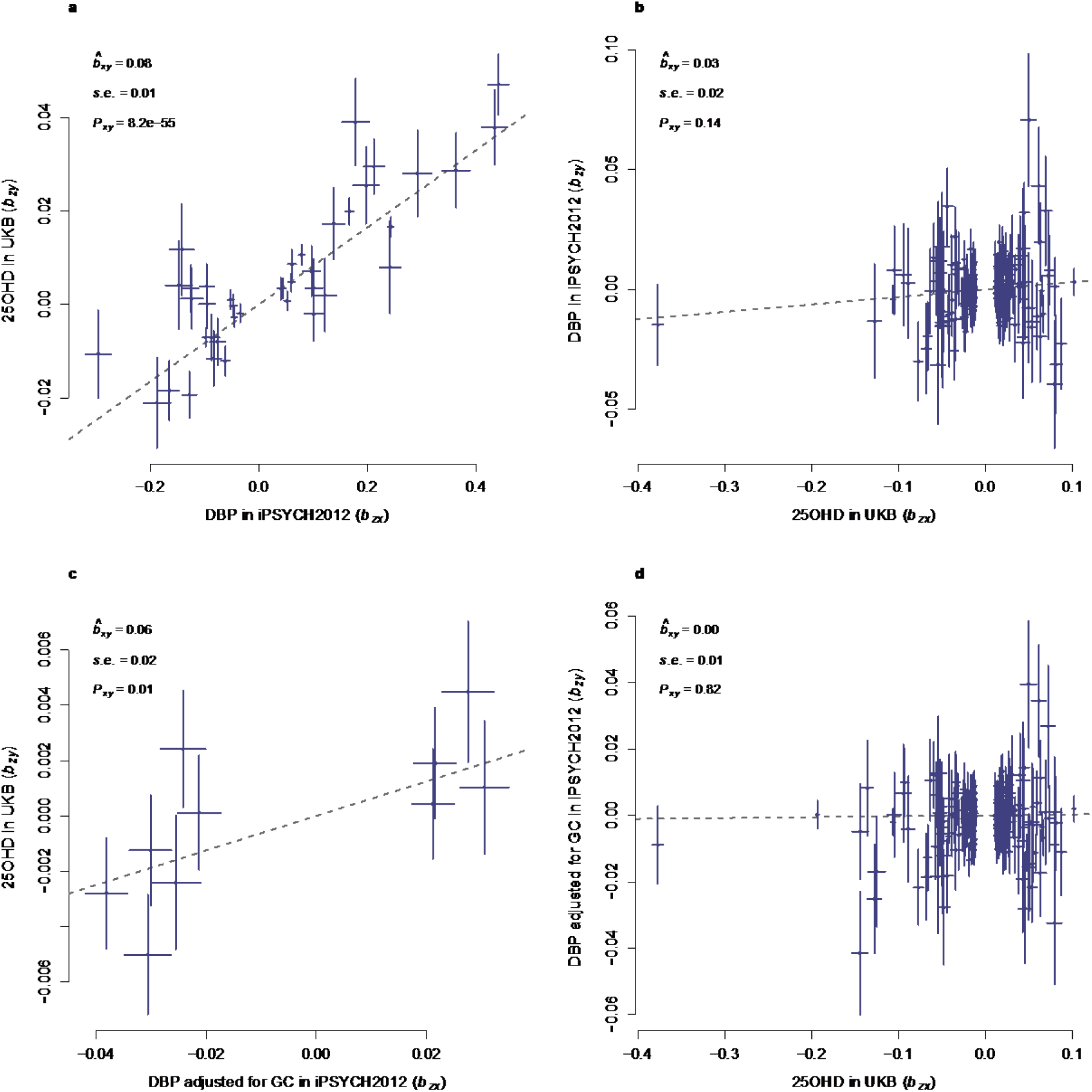
Causal effect of DBP on 25OHD. Shown in the top 2 panels are the GSMR results of DBP concentration, a) the effect of DBP on 25OHD and b) the effect of 25OHD on DBP. The GSMR estimates are shown in each panel. These GSMR estimates include estimate of effect, SE and *P*-value. The dash line represents GSMR estimate of effect. The summary statistics of DBP is from this study, conducted in iPSYCH2012. The summary statistics of 25OHD is from the study of Revez *et al*., conducted in UKB. Shown in the bottom 2 panels are the GSMR results of DBP concentration adjusted for GC genotypes, c) the effect of DBP adjusted for GC on 25OHD and d) the effect of 25OHD on DBP adjusted for GC. The GWAS of adjusted DBP was conducted with fitting the GC diplotypes as covariates. The summary statistics of 25OHD are the same as above, from the study of Revez *et al*. All the GSMR analyses were conducted with HEIDI-outlier. The SNPs which were identified as pleiotropy were excluded. The bar shown in the graph represents the GWAS SE at each SNP.

### GSMR relationships with other traits

There were no significant associations based on forward or reverse GSMR between GWAS summary statistics based on either DBP or DBP_GC versus any of the neuropsychiatric disorders. With respect to forward GSMR based on summary statistics based on DBP, there were no significant findings for any of the phenotypes.

With respect to forward GSMR based on GWAS summary statistics for DBP_GC, we found evidence to support causal associations between this phenotype and two autoimmune disorders (Figure 6a and 6b). First, we found a negative (i.e., protective) association between DBP_GC and multiple sclerosis (logOR = 0.65, SE = 0.18, *P*-value = 1.9×10^-4^, *N*SNPs = 13). Second, there was a negative association between DBP_GC and rheumatoid arthritis (logOR = 0.69, SE = 0.20, *P*-value = 7.4×10^-4^, *N*SNPs = 12). No pleiotropic SNPs were identified by HEIDI-outlier in either the multiple sclerosis or rheumatoid arthritis GSMR analyses.

**Figure 6.**
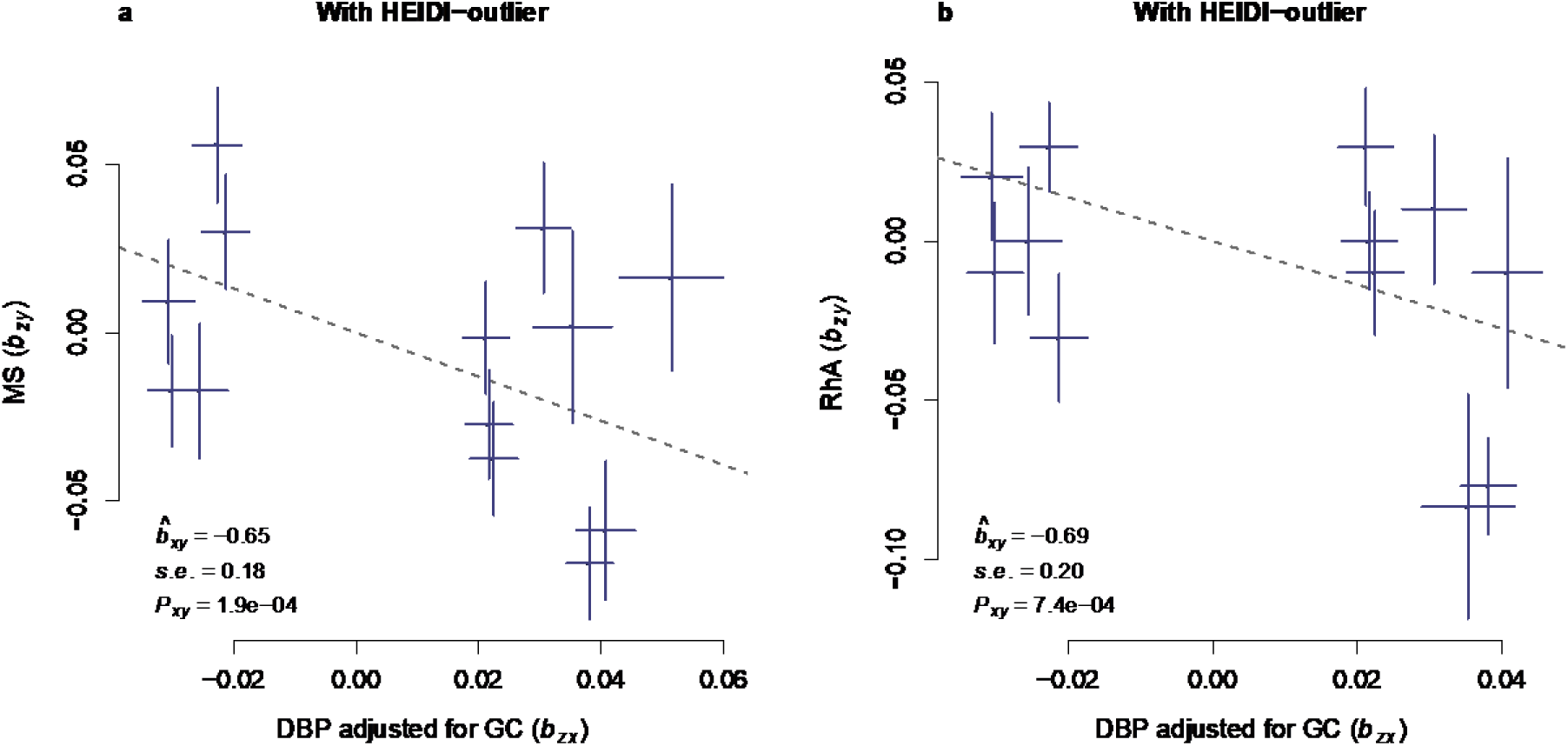
The GSMR effects of DBP concentration on multiple sclerosis (MS) and rheumatoid arthritis (RhA). The GSMR estimates are shown in each panel. These GSMR estimates include estimate of effect, SE and *P*-value. The dashed line represents the GSMR estimate of effect. No pleiotropic SNPs were identified by HEIDI-outlier for these two disorders. The bar shown in the graph represents the GWAS standard error for each SNP.

In addition, we found evidence to support the hypothesis that pleiotropic variants may influence the association between DBP_GC and two additional autoimmune disorders (Supplementary Figures S15a-d ). When potentially pleiotropic SNPs were removed, we found a positive association between DBP_GC and a higher risk of Crohn’s disease (logOR = 0.65, SE = 0.19, *P*-value = 7.6×10^-4^, *N*SNPs =11). This relationship was not apparent in analyses that included two potentially pleiotropic SNPs (rs11745587, rs56326707; logOR = -0.02, SE = 0.18, *P*-value = 0.28, *N*SNPs = 13). In general, the large positive GSMR estimate (when 2 pleiotropic SNPs were excluded) and the change in sign of the beta estimate when the 2 pleiotropic SNPs were included, suggests that we should be cautious in our interpretation of any potential association between DBP concentration and Crohn’s disease.

Furthermore, we identified a negative association between DBP_GC and risk of Type 1 diabetes when assessed without HEIDI-outlier test (logOR = -0.95, SE = 0.17, *P*-value = 1.2×10^-8^, *N*SNPs = 13 SNPs). When one pleiotropic SNP was removed from the analysis, the association became non-significant (logOR = 0.36, SE = 0.19, *P*-value = 0.06, *N*SNPs = 12 SNPs). The identified pleiotropic SNP was rs3184504, a missense variant in *SH2B3*. For both the findings related to Crohn’s disease and Type 1 diabetes, the opposite direction of beta coefficients in the presence or absence of potentially pleiotropic variants weakens the hypothesis that there is a direct influence of DBP_GC on these two disorders. Full details of the GSMR analyses are shown in Supplementary Data SD13 and SD14.

### PheWAS findings

Finally, we examined the association between the two DBP-related summary statistics and phenotypes in the UKB. For the GWAS summary statistics based on DBP, only one finding was significant (after Bonferroni adjustment for multiple comparison) (Supplementary Data SD15). We found a highly significant association with measured 25OHD concentration (part of the UKB biomarker set - Field ID: 30890; beta = 0.09, SE = 0.002, p <10E-100, *N* = 317,064). Note that the positive effect size (beta value) indicates that variants associated with higher DBP concentrations were associated with higher observed 25OHD concentration in UKB. Reassuringly, the association with clinical diagnosis of ‘vitamin D deficiency’ (ICD-10 E55; cases = 3150, noncases = 344,619) was negative and nominally significant (i.e. higher DBP associated with a reduced risk of a clinical diagnosis of vitamin D deficiency; beta = - 0.07, standard error = 0.02, p <0.001).

We dichotomized the continuous measure of 25OHD concentration in the UKB according to the Institute of Medicine definition of vitamin D deficiency (i.e. < 25 nmol/L)^52^. We then divided the PRS for DBP into 11 bins (quantiles). The 6^th^ (central) bin was set as the reference category. We tested the odds ratio of each binned PRS to the reference bin with logistic regression adjusted for sex, age and 20 PCs. Compared to the reference category, those in each of the 5 upper quantiles had significantly reduced odds of having vitamin D deficiency, while those in the two lowest quantiles had a significantly increased odds of having vitamin D deficiency (Figure 7; Supplementary data SD16). Compared to the highest quantile, those in lowest quantile has a 57% increased odds of having vitamin D deficiency (odd ratio = 1.57, 95% confidence intervals 1.49-1.65).

**Figure 7:**
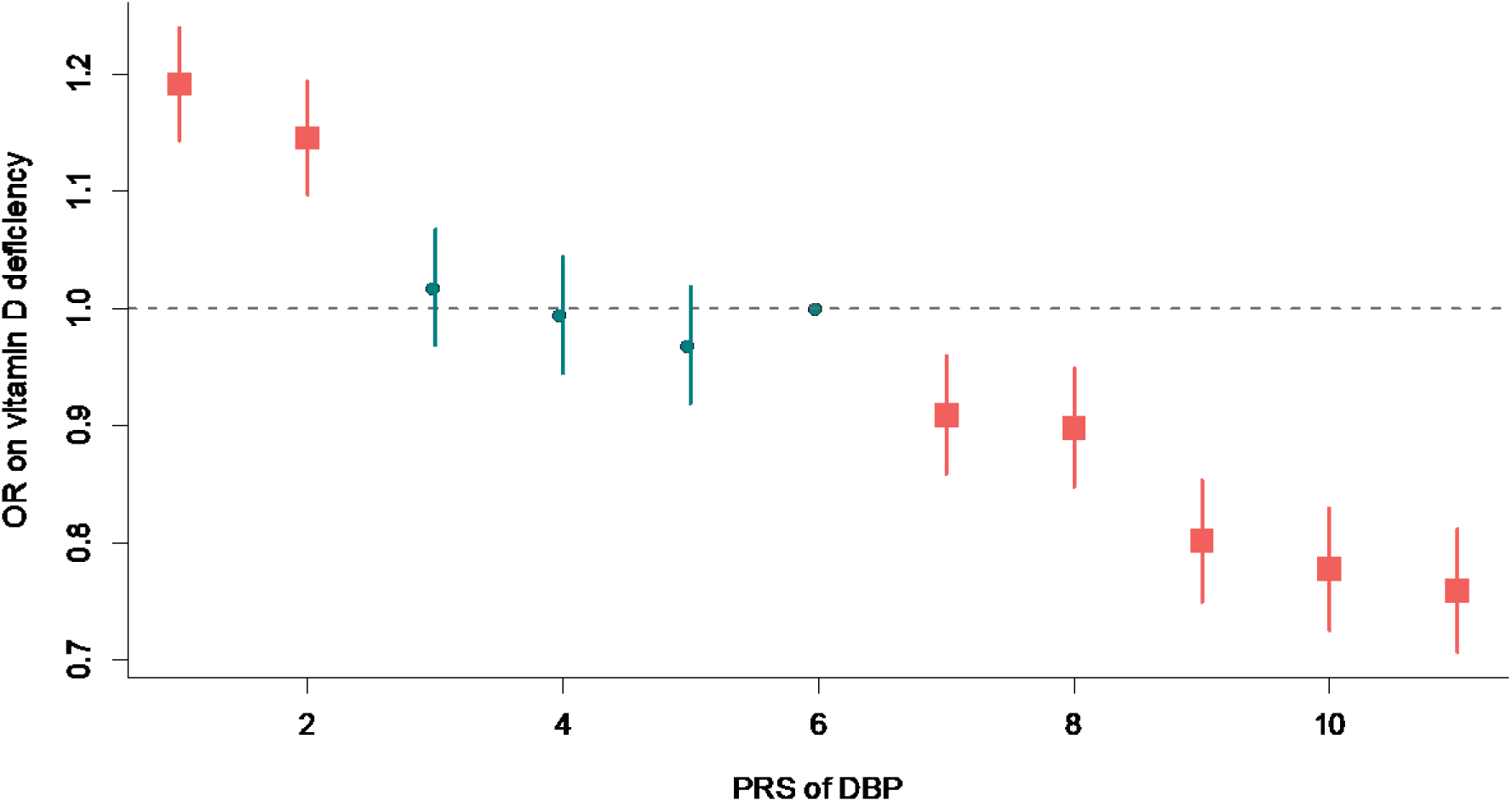
Odds ratio (OR) of DBP PRS on vitamin D deficiency. The PRS of DBP was divided into 11 bins. The 6^th^ bin with the average of PRS is set as the reference category (odds ratio = 1). The plot shows the odds ratio of PRS for each bin compared to the reference bin. The vertical bars represent the 95% confident interval. The red colour represents the significant estimates, with *P*-value < 0.05.

For the DBP_GC DBP summary statistics, we found a range of associated phenotypes (Supplementary Data SD17). For example, summary statistics based on DBP_GC summary statistics were protective for the clinical diagnosis of essential (primary) hypertension (logOR = -0.03, SE = 0.004, *P*-value = 2.52×10^-11^; *N*cases = 104,892, *N*non-cases = 242,876).

Consistent with this finding, DBP_GC was also negatively associated with both observed diastolic and systolic blood pressure in a large UKB subsample (*N* ∼ 253,370, *b*diastolic = -0.02, SE = 0.002, *P*-value = 1.7×10^-23^; *b*systolic = -0.01, SE = 0.002, *P*-value = 5.1×10^-11^ respectively). In addition, DBP_GC was associated with a) reduced pulse rate (*b* = -0.01, SE = 0.002, *P*-value = 8.33E-0.09; *N* = 324,967), (b) gastritis and duodenitis (logOR = -0.03, SE = 0.01, *P*-value = 7.56×10^-7^; *N*cases = 39,620, *N*non-cases = 308,147); and associated with an increased risk of (c) vasomotor and allergic rhinitis (logOR = 0.03, SE = 0.01, *P*-value = 6.08×10^-6^; *N*cases = 34,276, *N*non-cases = 313,476) and (d) agranulocytosis (logOR = 0.06, SE = 0.01, *P*-value = 1.49×10^-5^; *N*cases = 4,919, *N*non-cases = 342,850). There was no association between higher DBP_GC and observed 25OHD concentration (*b* = 0.003, SE = 0.002, *P*-value = 0.11; *N* = 317,064), but again we found a nominally significant association with a prior clinical diagnosis of vitamin D deficiency (logOR = -0.04, SE = 0.02, *P*-value = 0.02; *N*cases = 3,150, *N*non-cases = 344,619).

Finally, DBP_GC was associated with (a) an education-related measure (higher concentration of DBP associated with more years in education), (b) birthweight (higher concentration of DBP associated with higher birthweight) and (c) two adult anthropometric measures (higher concentration of DBP associated with reduced body fat percentage).

PheWAS plots for both the DBP and DBP_GC based analyses can be found in Supplementary Figures S16 and S17 respectively.

## DISCUSSION

We identified 26 independent loci associated with DBP concentration, 24 of which were either in or in close proximity to the *GC* gene. When we adjusted for key *GC* haplotypes, we identified 15 loci distributed over 10 chromosomes. We confirm the robust influence of *GC*- related variants on the concentration of DBP, and provide new clues as to the genetic complexity of this highly polymorphic protein. Mendelian randomization suggests that variants related to increased DBP concentration are causally related to higher 25OHD concentration, but not vice versa. Our findings related to autoimmune disorders were of particular interest — Mendelian randomization analyses lend weight to the hypotheses that DBP-related mechanisms influence the risk multiple sclerosis and rheumatoid arthritis. The following discussion focuses on six key findings.

First, we confirm that the genetic architecture of DBP concentration is characterized by highly influential loci within or near the *GC* gene. In particular, over half (52.6%) of the variance in DBP concentration is explained by two canonical missense variants (rs7041 and rs4588). Consistent with previous literature, we found that the proportions of the different haplotypes varied by genetically-defined ancestry (Supplementary Figure S10). While each of the six key *GC*-related diplotypes were detected within the group defined as European ancestry, DBP concentrations still showed appreciable variation *within* each of these groups. The genetic correlates underpinning this additional variation were foregrounded in the *GC*- adjusted GWAS, which identified 15 COJO independent loci distributed over 10 chromosomes (only two were on chromosome 4, in proximity to *GC*).

Second, we show that DBP is highly heritable. Using related individuals, the narrow-sense heritability was 68%. The estimate is similar to the heritability (60%) reported by Moy and colleagues^9^. When we examined how much of the variance in DBP concentration could be attributed to common single nucleotide variants included in the GWAS, the proportions remained appreciable, 53% for the *GC* gene and 5% for the remaining genotypes. The results were consistent across the three methods with different assumptions of genetic architecture, GCTA-GREML, SBayesS and LDpred2-auto. Moreover, we identified 15 independent COJO loci after the adjustment for the *GC* haplotypes. These 15 COJO SNPs explained 0.49% variance of DBP in total. The remaining ∼4.5% (5% - 0.49%) variance is likely to be captured by SNPs which were not significant in the current GWAS. These findings suggest DBP has polygenic features, in addition to the very large genetic variance encoded by the *GC* gene. These findings reinforce the value of the *GC*-adjusted GWAS and related post-GWAS analyses.

Third, based on our sample of ∼ 65,000 European- ancestry individuals, we found that the genetic architecture of 25OHD in neonates was consistent with that reported by similar-sized GWAS studies based on adults^10, 12^. We found one quasi-independent locus in the *GC* gene. Furthermore, based on the correlation between the effect sizes for the SNPs identified in the UKB-based GWAS (n ∼ 350,000 adults)^11^ and the subset of these SNPs available in our neonatal sample, a significant positive association was found (Pearson *r* = 0.66, *P*-value < 2.2×10^-16^). The family-based heritability for 25OHD was comparable to that reported by Revez et al. in the UKB sample^11^ (current study = 36%; UKB = 32%). It is important to note that neonatal 25OHD concentration is entirely reliant on maternal 25OHD concentrations^53^ and while the correlation between the maternal and offspring genotypes would be 0.5, the genetic correlates of neonatal 25OHD may be more strongly correlated with the (unobserved) maternal genotype, rather than the (observed) neonatal genotype. Because maternal DBP does not cross the placenta^1^, this is not an issue when examining the genetic correlates of DBP in neonates.

Fourth, we identify new candidate loci that influence DBP concentration. As expected with a highly polymorphic protein like DBP, many of the quasi-independent loci (17 of 26 in the main analysis) were in or very close to the *GC* gene (including the canonical missense variant rs7041). Fine-mapping identified: (a) a missense variant in *SH2B3* (rs3184504), which encodes a widely-expressed protein involved in the activation of kinase signaling activities, and which has been linked to a range of disorders, including diabetes^54^, and (b) a missense variant in *GSDMA* (rs56030650) which encodes a precursor of a pore-forming protein that can influence membrane permeabilization. Variants in this gene have been linked to pyroptosis (inflammatory cell death) and inflammatory bowel disease.^55^

Fifth, our findings provide convergent evidence that variants that influence DBP concentration influence 25OHD concentration, but not vice versa. Apart from the findings from Mendelian randomization, we found no significant variation in DBP concentration by month of testing (in contrast to 25OHD, which shows marked seasonal variation). The PheWAS results confirm that variants related to higher DBP concentration are associated with (a) higher concentration of 25OHD and (b) a reduced risk of receiving the clinical diagnosis of vitamin D deficiency. In light of evidence from clinical trials indicating that vitamin D supplementation does not impact on DBP concentration^56^, our findings lend additional support to the unidirectional nature of the relationship between DBP and 25OHD.

It has long been appreciated that: (a) variants in the *GC* gene influence the concentration of DBP, and (b) that variants within the *GC* gene are robustly and consistently associated with 25OHD concentration. Over the last few decades there has been a focus on the relationship between (a) total 25OHD (i.e., the value routinely measured by laboratories measures both protein-bound and free 25OHD), and (b) free 25OHD (directly observed by specialized assays or estimated based on prediction models)^2,57^. It is clear that free 25OHD concentration is strongly correlated with the total 25OHD concentration^7^. Our Mendelian randomization findings lend weight to the hypothesis that the concentration of DBP is causally related to the concentration of (total) 25OHD. In light of (a) recent clinical evidence from individuals with homozygous deletions or pathogenic variants of the *GC* gene^5,58^, and (b) findings from *GC* knock-out animal models^6^, the concentration of DBP has been proposed to be a key factor in determining the half-life of vitamin D metabolites, as unbound 25OHD is more rapidly transferred to target cells and catabolised^5,7^. By extension, those with lower concentration of DBP would be more likely to experience vitamin D deficiency, because this would shorten the functional half-life of 25OHD. A study tracking the excretion of deuterium-labelled 25OHD supports this hypothesis.^59^ If two individuals have identical concentration of total 25OHD (free and bound) at baseline, then in the absence of new vitamin D production, over a given period the individual with higher concentration of DBP would be less likely to subsequently develop vitamin D deficiency because of the longer half-life of 25OHD. Higher DBP concentration can act as a larger reservoir for 25OHD, and thus provide a more effective ‘buffer’ against future vitamin D deficiency. We speculate that (a) observed lower DBP concentration, and/or (b) lower polygene risk scores based on summary scores from DBS-related GWASs, may provide an informative proxy measure related to an increased future risk of vitamin D deficiency.

Sixth, we provide new evidence from Mendelian randomization that links DBP concentration and risk of several autoimmune disorders. There is already a strong body of literature based on observational epidemiology and Mendelian randomization linking low vitamin D status and an increased risk of multiple sclerosis^17, 60–65^. Based on Mendelian randomization, we also found that increased DBP concentration was associated with a reduced risk of rheumatoid arthritis. In keeping with our findings related to multiple sclerosis, the effect size of this association was substantial (logOR = -0.65, SE = 0.18, *P*-value = 1.9×10^-4^). There is evidence linking vitamin D deficiency and an increased risk of rheumatoid arthritis^66, 67^.

The active form of vitamin D (1,25OHD) is an immuno-modulator, and has anti-inflammatory effects^68^. Because we identified these auto-immune-related findings only in the summary statistics generated from the DBP_GC analysis (which is a weaker instrument for 25OHD concentration compared to the unadjusted DBP), this raises the possibility that these findings may reflect non-vitamin D associated properties of DBP (e.g., C5a-mediated chemotaxis, T-cell response, macrophage activation^1,69^). It could also be argued that the established links between 25OHD concentration and risk of several autoimmune disorders provides a more parsimonious explanation for these particular findings. We hope that our findings can stimulate hypothesis-driven research focused on the role of DBP for multiple sclerosis and rheumatoid arthritis.

Our study has several strengths. Our sample was over 30 times larger than the only other published GWAS of DBP^9^. We were able to assess 25OHD in the same large sample, and also confirm our findings within a subcohort representative of the general population. Our findings also have several important limitations. The sample were neonates at the time of testing, and it remains to be seen if the genetic architecture of DBP identified in our study will generalize to adult populations. We know that certain factors (e.g., pregnancy, use of the oral contraceptive pill) and several disorders that lead to proteinuria can impact on DBP concentration in adults^70^. For the assessment of DBP, we used a monoclonal antibody pair, which resulted in a similar bias towards the 1S-isoform of DBP as previously reported in studies comparing Americans of African and Caucasian origin (which also used immunoassays based on monoclonal antibodies) ^8,71–74^. However, we restricted the sample to those with European ancestries and conducted the GWAS with and without adjustment for GC haplotypes. Our sample is enriched with people with mental disorders; however, we found no forward or reverse GSMR association between mental disorders and DBP. In addition, we did a planned sensitivity analyses where we ran the GWAS again only in the population-based subcohort (a rare resource that is free of ascertainment bias). The GWAS results in the full case-cohort versus the sub-cohort sample were comparable. In addition, because our main analyses were restricted to Europeans, there is a need to examine the genetic correlates of DBP concentration in more diverse ancestry groups. Finally, the DBP_GC analysis also identified a SNP (rs635634) that is ∼4 kb upstream from the *ABO* gene. We noticed that recent studies of the genetic correlates of the human plasma proteome have identified genes that encode for ‘master regulator’ proteins – variants in these genes have widespread correlations with the concentration of other circulating proteins^75, 76^.

*SH2B3* and *ABO* were both identified as having associations with over 50 other protein concentrations. However, variants in these same genes have also been associated with hematocrit^77^, which may have downstream consequences on the accuracy of a range of sera-based^78^ and dried blood spot-based assays^79^. The influence of genetic variants related to hematocrit on (a) the accurate quantification of blood-based biomarkers, and (b) subsequent GWASs based on these measurements, warrants additional investigation.

Our findings may have clinical consequences – those with genetic variants associated with lower DBP concentrations may have a particular requirement for vitamin D supplementation over the course of winter (compared to those with genetic variants associated with higher DBP concentrations). If our Mendelian randomization findings related to multiple sclerosis and rheumatoid arthritis are replicated in future studies, there may be a case to ensure that those with a genetic predisposition to lower DBP concentrations are encouraged to take regular vitamin D supplements. We note with interest that a recent large randomized controlled trial found that vitamin D supplementation reduced the incidence of several autoimmune disorders^16^. If previously completed randomized controlled trials of vitamin D supplementation have access to the genotype of their participants, we speculate the use of supplements may be associated with superior outcomes in those with lower genetically-predicted DBP concentration.

## Data Availability

Owing to the sensitive nature of these data, individual level data can be accessed only through secure servers where download of individual level information is prohibited.  Each scientific project must be approved before initiation, and approval is granted to a specific Danish research institution. International researchers may gain data access through collaboration with a Danish research institution. More information about getting access to the iPSYCH data can be obtained at https://ipsych.au.dk/about-ipsych/.

## Acknowledgments

The Genotype-Tissue Expression (GTEx) Project was supported by the Common Fund of the Office of the Director of the National Institutes of Health, and by NCI, NHGRI, NHLBI, NIDA, NIMH, and NINDS. The data used for the analyses described in this manuscript were obtained from the GTEx Portal on 02/01/22.

The authors thank GenomeDK and Aarhus University for providing computational resources and support that contributed to these research results. This research has been conducted using the UK Biobank Resource under Application Number 12505.

## Authors’ contributions

Laboratory 25OHD and DBP assays: SGB, ASC, NBL, KS.

Genetic analyses: CA, ZZ, BJV

Data interpretation and manuscript preparation: CA, ZZ, BJV, JMcG.

iPSYCH sample funding, design and management: DH, ADB, TW, MN, PBM.

ANGI sample funding, design, management, analysis: CMB, LVP

Manuscript editing and final approval: all authors

## SUPPLEMENTARY MATERIAL

### SUPPLEMENTARY METHODS

#### Methods S1 Sample preparation steps

Two 3.2 mm disks from neonatal dried blood spot (DBS) samples were punched into each well of polymerase chain reaction plates (72.1981.202, Sarstedt). 130 µL extraction buffer (PBS containing 1% BSA (Sigma Aldrich #A4503), 0.5% Tween-20 (#8.22184.0500, Merck Millipore), and complete protease inhibitor cocktail (#11836145001, Roche Diagnostics)) was added to each well, and the samples were incubated for 1 hour at room temperature on a microwell shaker set at 900 rpm. After separating the extract from the filter paper into sterile Matrix 2D tubes (#3232, ThermoFisher Scientific), the extracts were stored at -80°C for 6-7 years before analysis.

#### Methods S2 DBP concentration and pre-analytical variation

To validate the stability of the samples during storage, we randomly selected 15-16 samples from five years (1984, 1992, 2000, 2008, and 2016; a total of 76 samples). After extracting the samples and adding them to a MSD plate, the rest of the extracts were frozen for 2 months, thawed and measured as described above to imitate the freeze–thaw cycle of the samples in the study. This showed (Figure S1 – data from samples that were frozen for 2 months prior to analysis) that samples from 1984 had higher concentration compared to more recent samples. A likely explanation for this variation would be the change of filter paper in 1989, where “Schleicher & Schuell grade 2992” was replaced by “Schleicher & Schuell grade 903”.

#### Methods S3 Assay of 25OHD concentration

For the assay of 25OHD, 30 µL of each sample was transferred to a Thermo Scientific 96 well polypropylene storage microplates before 120 µL internal standard (reconstituted in acetonitrile and diluted to a working solution of 1:100 compared to the kit insert) was added. After centrifugation the samples were prepared for a liquid-liquid extraction procedure. 200 µL of the upper organic phase (containing the purified vitamin D metabolites) was transferred to a Thermo Scientific^TM^ WebSeal Plate+ 96-Well Glass-Coated Microplate. The samples were dried down in an Eppendorf Bench Top Concentrator Plus^TM^ (60°C) before the vitamin D metabolites were derivatized with 20 µL of the commercial PTAD reagent (reconstituted in ethyl acetate and diluted to a working solution of 1:12). After incubation and quenching (by the addition of 50 µL ethanol), samples were dried down in a concentrator before being reconstituted in 80 µL 1:1 acetonitrile/deionized water solution. After reconstitution, 40 µL was injected into the LC-MS/MS system. The LC system is a Thermo TLX2 Turboflow system, comprised of a CTC Analytics HTS PAL autosampler, a dual LC system (one Agilent 1200 quartinary and one Agilent 1200 binary pump) and two Thermo Scientific hot pocket column heaters. The LC systems are interfaced with a triple quadrupole mass spectrometer (Thermo Scientific TSQ Quantiva) equipped with a heated electrospray ionization probe. The LC system is controlled by Aria MX Direct Control software, whereas the mass spectrometer is controlled in the TSQ Quantiva Tune Application software (version 2.0.1292.15). Thermo TraceFinder^TM^ 3.2 application software is used to acquire and process data.

Intermediate precision was obtained by quantifying the concentration of three stable-isotope labeled external commercial quality controls with a low, medium and high concentration of each vitamin D metabolite. The percent coefficient of variances ranged from 4.7-7.2%. With respect to the accuracy of the method, SRM^®^ 972 sera from the National Institute of Standards and Technology (NIST) were mixed 1:1 with purified erythrocytes and spotted onto filter paper before the proteins were extracted followed by the extraction, derivatization and quantification of the endogenous levels of 25OHD_2_ and 25OHD_3_ as described above. The evaluation of the SRM^®^ 972 from NIST demonstrated an excellent accuracy (92-105%). The method was able to detect a concentration of both 25OHD_2_ and 25OHD_3_ down to approximately 5 nmol/L in the protein extracts. The laboratory participated in the Vitamin D External Assessment Scheme (DEQAS)^1^ during the period of the study (2018-2021) with a mean (range) bias from the target values of 3.8% (-10.6, 12.6).

#### Methods S4 Genetic ancestry inference

By design, the iPSYCH case-cohort samples are born in Denmark. To infer their genetic ancestry we used the sample’s parental country of birth as proxy, as determined from the Danish Registers. First, we identified the subset of individuals in which both parents were born in the same region (“Africa”, “Asia”, “Australia”, “Denmark”, “Europe”, “Greenland”, “Middle East”, “N.America”, “S.America”, “Scandinavia”). The regions “Denmark”, “Europe”, “N.America”, “S.America”, “Scandinavia”, “Australia” were all re-defined as “Europe”. We then looked at country of birth of the father and kept only countries where there were > 10 individuals born in that country.

Using the father’s country of birth as the grouping variable, we calculated the geometric median of the first 20 PCs per country. Then we calculated the distance to all country centers and applied a hierarchical clustering algorithm (base r hclust function with method = “single”). The population centers were then chosen based on a visual inspection of the clusters as the country with the largest sample size. The following countries were chosen as population centers: “Turkey”, “Kingdom of Morocco”, “Islamic Republic of Pakistan”, “Denmark”, “Somali Republic”, “Socialist Republic of Vietnam”, “The Gambia”. After choosing the cluster centers, all other samples were assigned to the nearest cluster inside a threshold defined as: thr_sq_dist = 0.002 * (max(dist(all_centers)^2) / 0.40) (Supplementary Figure S2). The cluster tags were changed from country names to geographical region names, as individuals from nearby countries where clustered together in the final classification. The PC1 vs. PC2 plot of the different ancestry clusters is shown in Supplementary Figure S3.

#### Methods S5 iPSYCH independent replication sample

From the European ancestry definition described above, we identified a replication sample of nearly- European individuals by expanding the threshold around the center of the European cluster to thr_sq_dist = 0.002 * (max(dist(all_centers)^2) / 0.10) (Supplementary Figure S4). This resulted in a sample of 1,881 individuals of nearly-European ancestry. From these we identified 1,529 individuals not related to each other or to anyone in the main analysis (i.e., all GRM off-diagonals <|0.05|).

Supplementary Figure S5 shows the PC1 vs. PC2 plot of the replication sample compared to the other ancestry clusters.

#### Methods S5 Summary statistics for target phenotypes used in GSMR analyses

We obtained summary statistics for the following phenotypes (see below for reference details): schizophrenia^2^, major depression^3^, bipolar disorder^4^ , autism spectrum disorder^5^, attention deficit hyperactivity disorder^6^ , Alzheimer’s disease^7^, educational attainment^8^, multiple sclerosis^9^, amyotrophic lateral sclerosis^10^, type 1 diabetes^11^, Crohn’s disease^12^, ulcerative colitis^12^ , rheumatoid arthritis^13^

### SUPPLEMENTARY TABLES

**Tables S1.** GC haplotypes Legend S1: The individual’s GC gene haplotypes were constructed based on the combination of the GC isoforms (not phased data). The GC haplotypes can be inferred from the specific allele combination of the two SNPs rs4588 and rs7041 as GC 1f (rs7041-T & rs4588-C), GC 1s (rs7041-G & rs4588-C) and GC 2 (rs7041-T & rs4588-A). Both the SNPs were hard-called genotyped for the iPSYCH data, as they are included in the PsychChip v1.0 array. Table S1 shows the coding of the SNPs in the chip.

| CHR | SNP | POS (GRCh37) | freq | A1 | A2 |
| --- | --- | --- | --- | --- | --- |
| 4 | rs4588 | 72618323 | 0.727 | G | T |
| 4 | rs7041 | 72618334 | 0.428 | A | C |

**Table S2.**
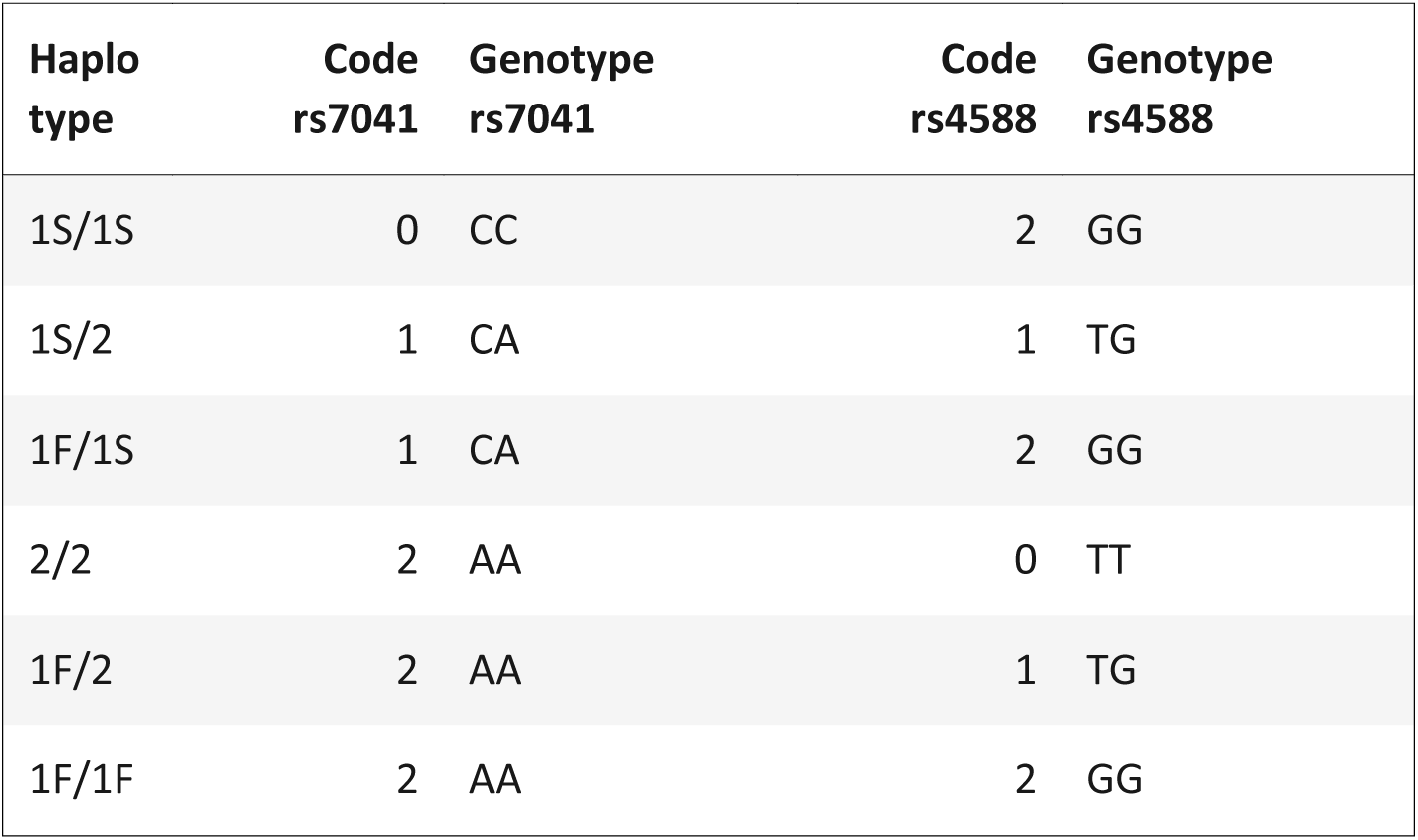
GC-based haplotype combinations (diplotypes). **Legend S2:** Both SNPs have been coded using the opposite strand to the isoform definition, but we defined the haplotypes based on the genotype call in iPSYCH. The code indicates the number of copies of the alternative allele (A1). The conversion between codes and the 6 GC haplotype combinations is shown in the table below. Ignoring chromosome phasing, this allows 6 possible diplotypes.

| Haplo<br>type | Code<br>rs7041 | Genotype<br>rs7041 | Code<br>rs4588 | Genotype<br>rs4588 |
| --- | --- | --- | --- | --- |
| 1S/1S | 0 | CC | 2 | GG |
| 1S/2 | 1 | CA | 1 | TG |
| 1F/1S | 1 | CA | 2 | GG |
| 2/2 | 2 | AA | 0 | TT |
| 1F/2 | 2 | AA | 1 | TG |
| 1F/1F | 2 | AA | 2 | GG |

### SUPPLEMENTARY FIGURES

**Figure S1.**
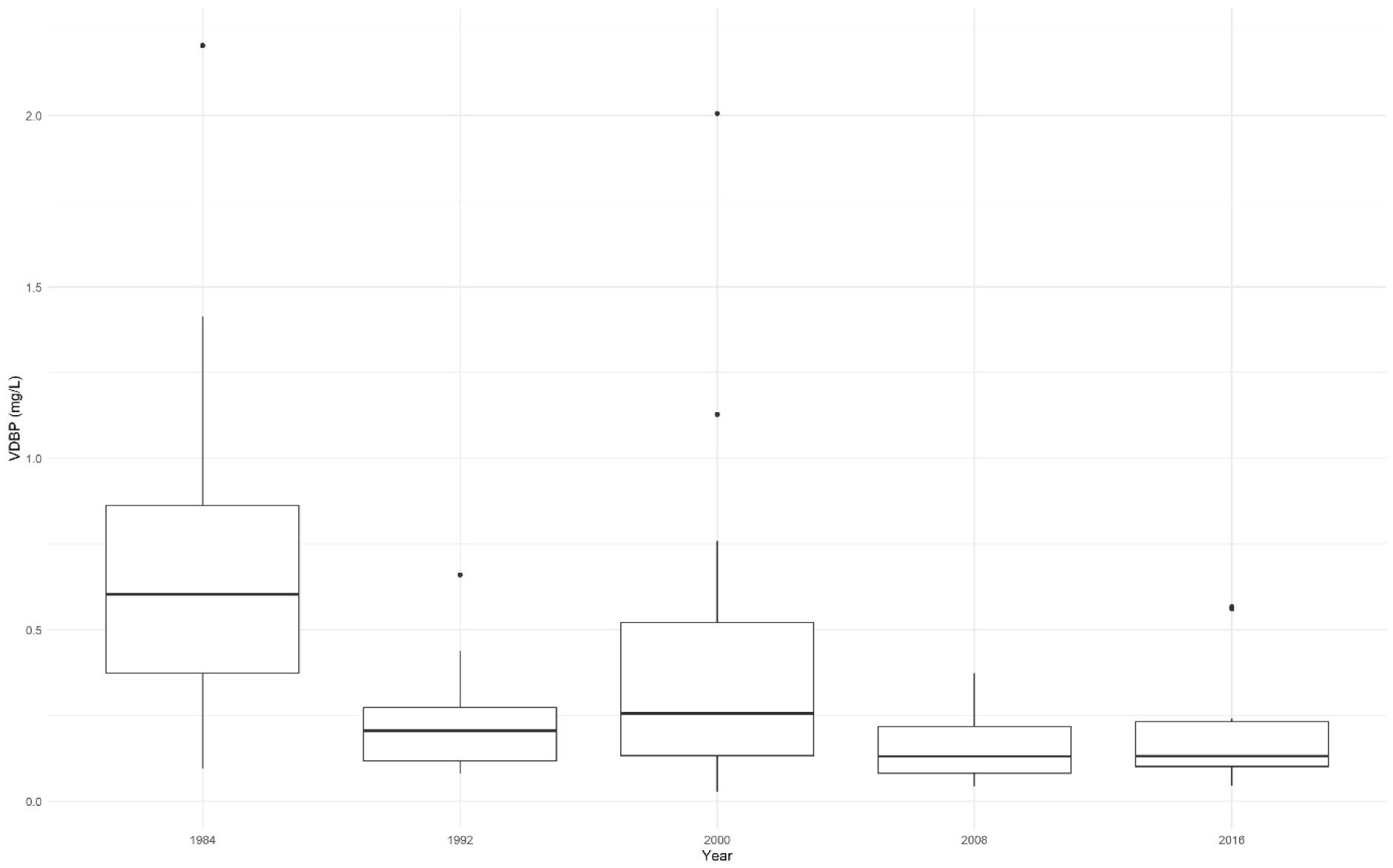
Boxplots showing the median value of DBP concentration split by year of birth Boxplots showing the median value of DBP concentration split by year of birth (x-axis). Y- axis displays concentration in mg/L. Hinges of boxes are first and third quartiles.

**Figure S2.**
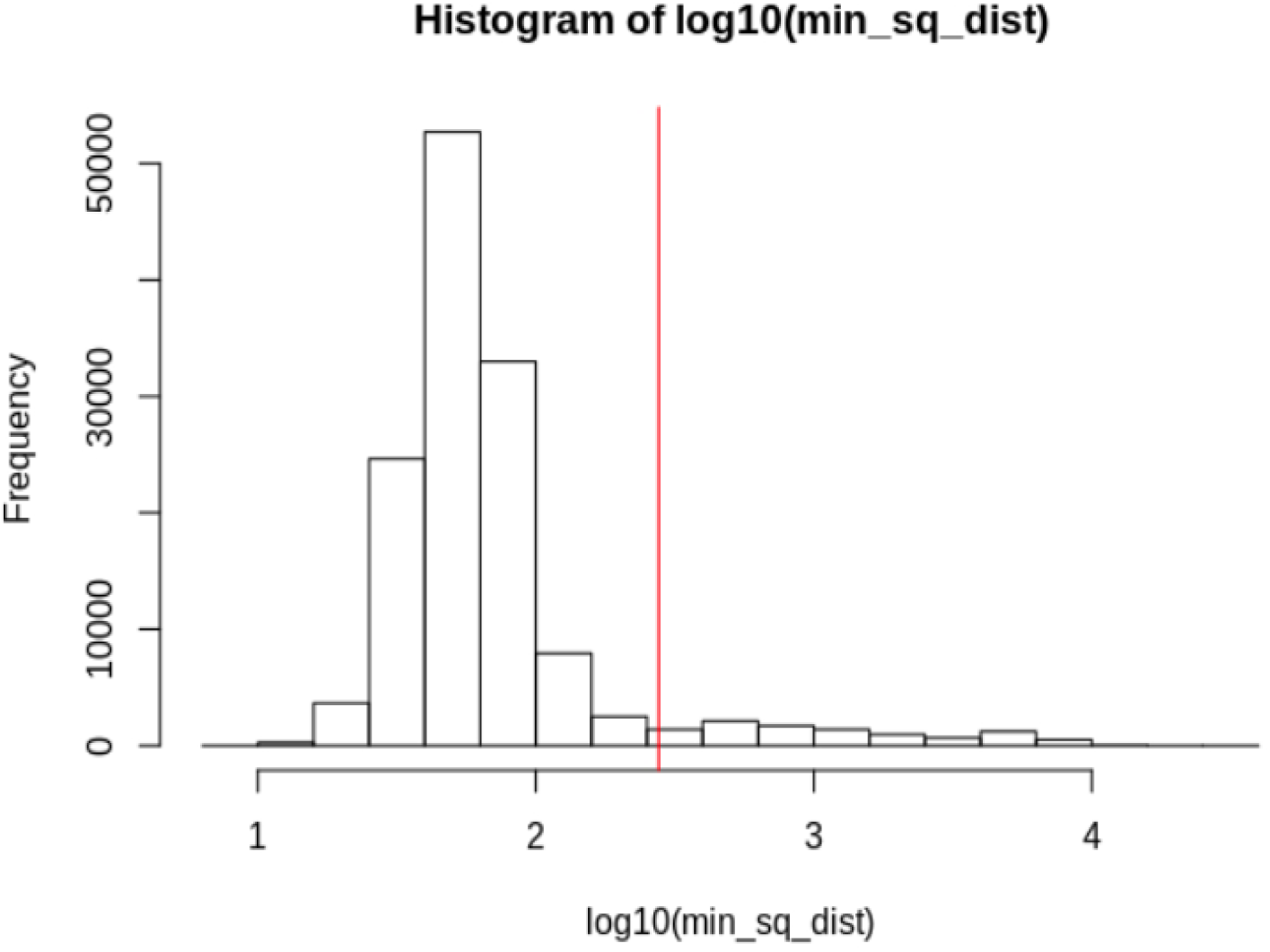
Genetic ancestry - Histogram of distance to cluster centre

**Figure S3.**
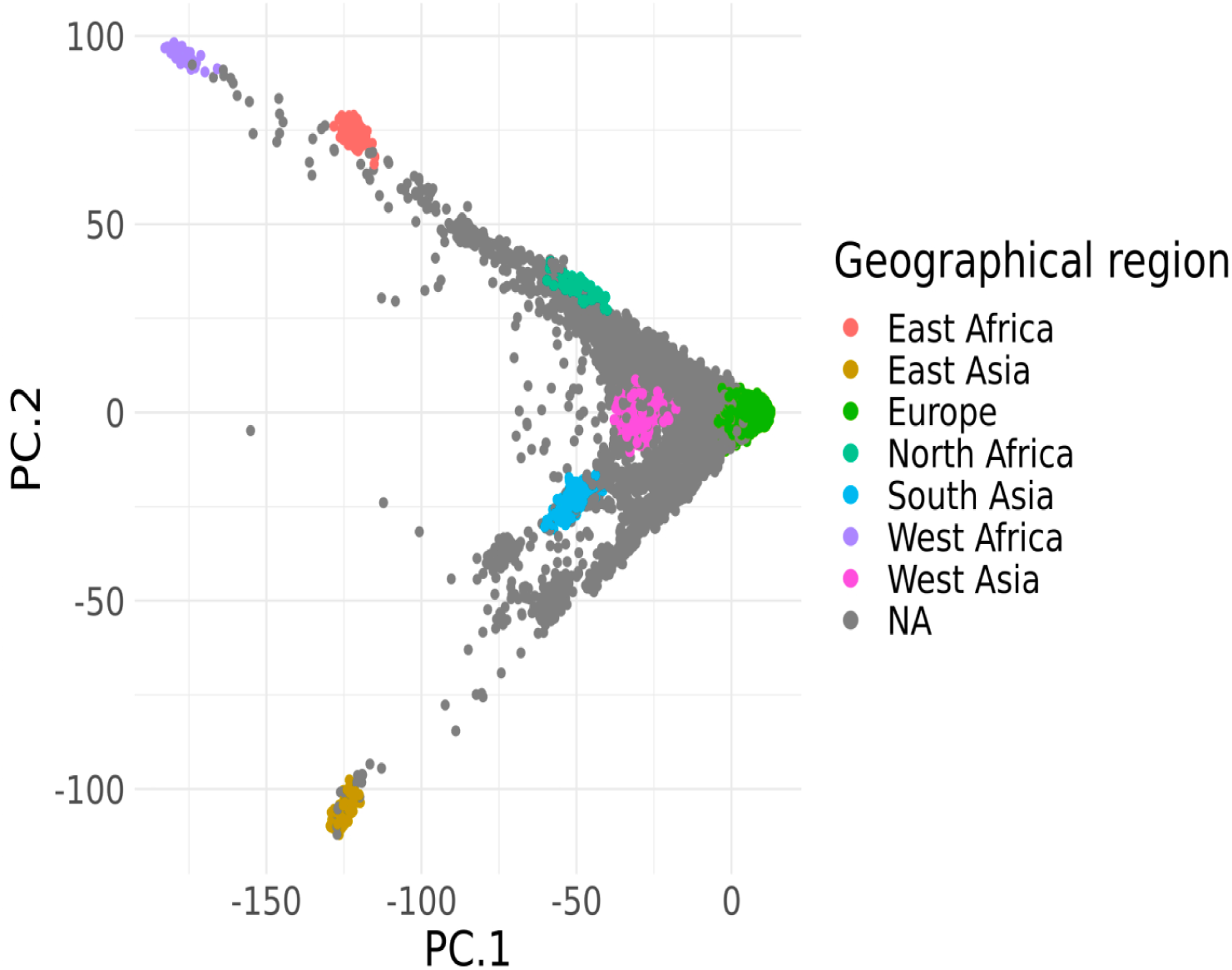
Genetic ancestry – plot of principal component 1 (PC1) versus principal component 2 (PC2)

**Figure S4.**
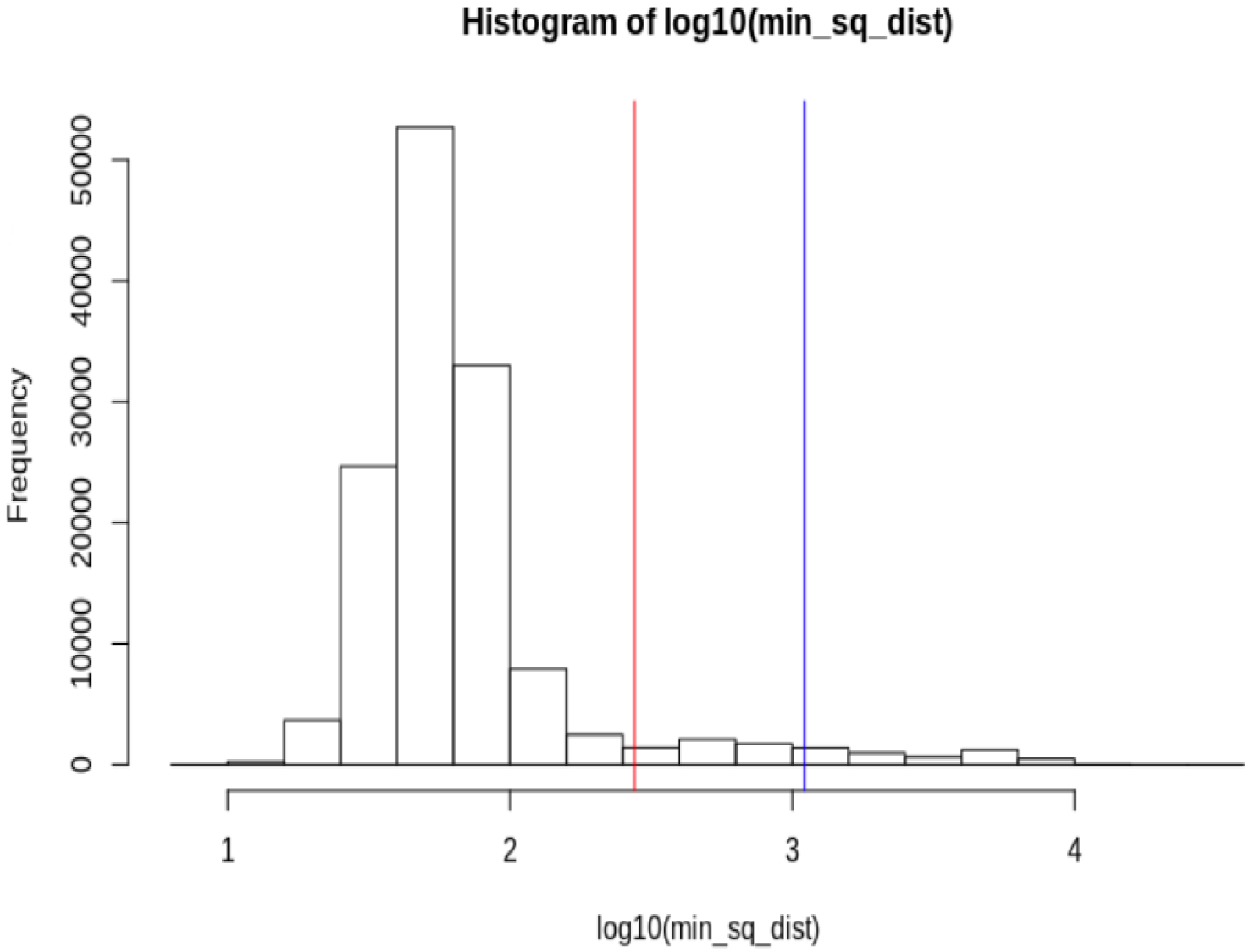
Defining the near-European replication sample – distance to centre cluster

**Figure S5.**
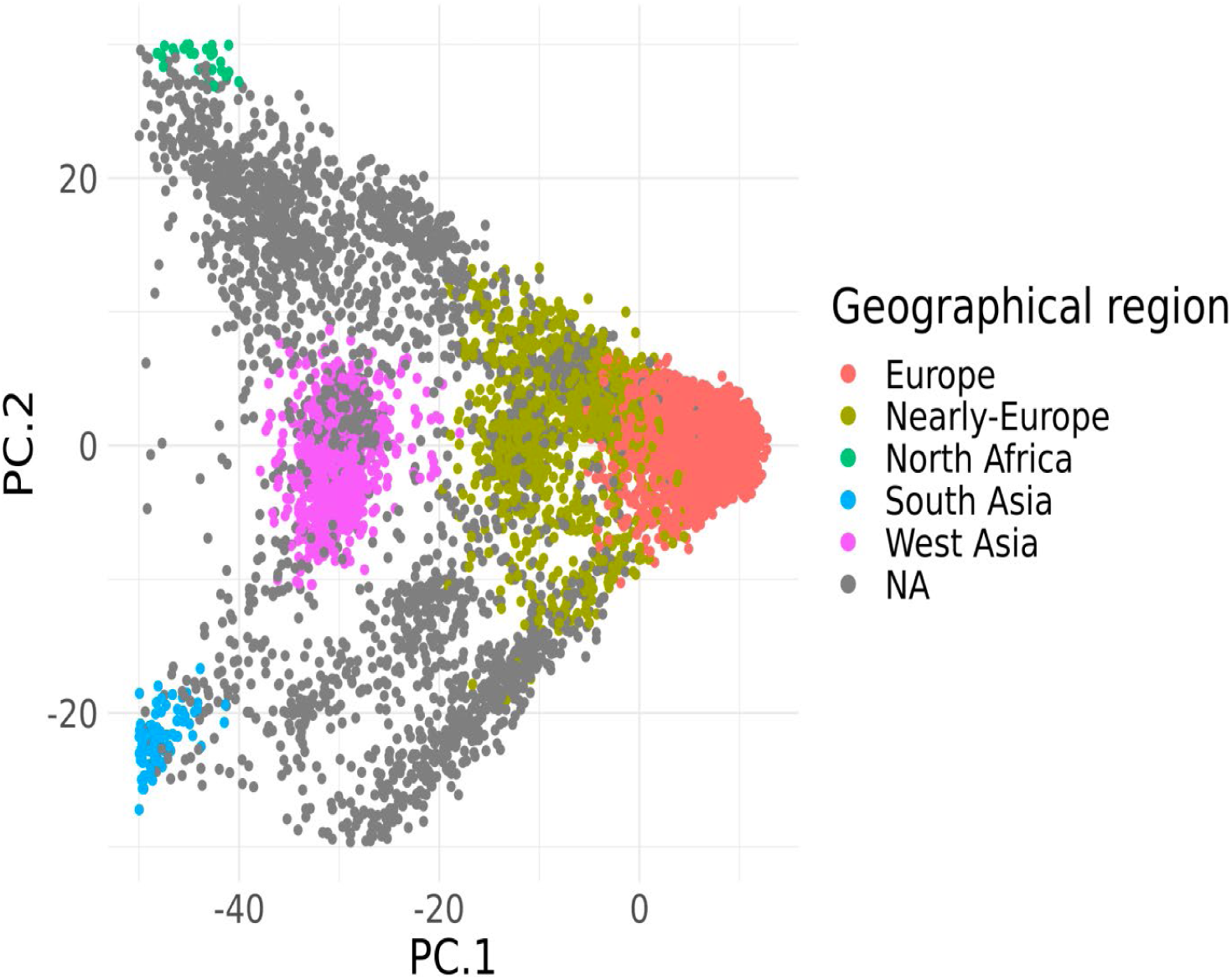
Defining the near-European replication sample – principal component 1 (PC1) versus principal component 2 (PC2)

**Figure S6.**
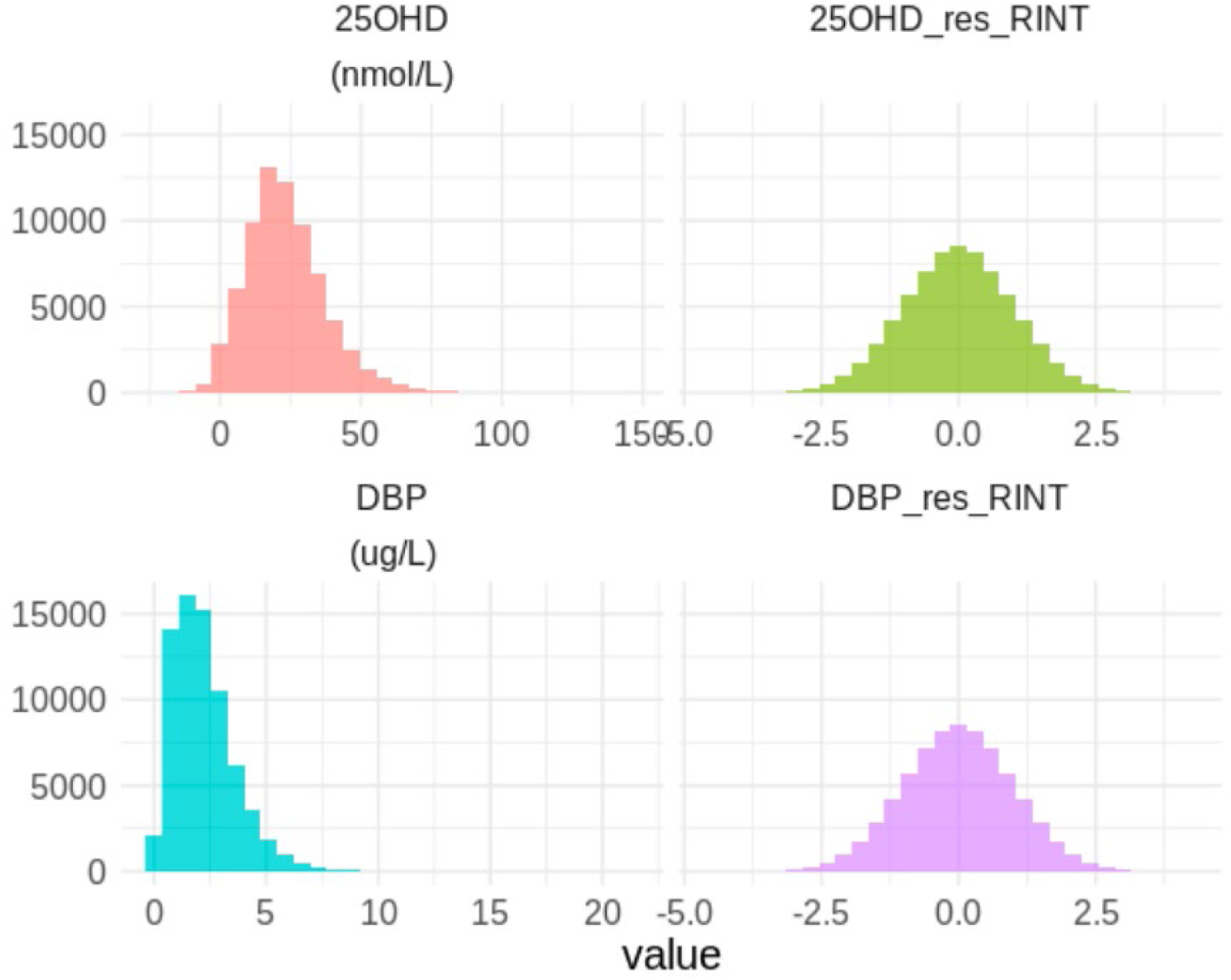
Distribution of 25OHD and DBP before and after transformation Raw and residualized-ranked inverse normal transformation (RINT) phenotypes. In the phenotypes suffixed with res_RINT, the quantification plate effect has previously been regressed out with a linear mixed model, and a rank-inverse normal transformation (RINT) applied to the residuals.

**Figure S7.**
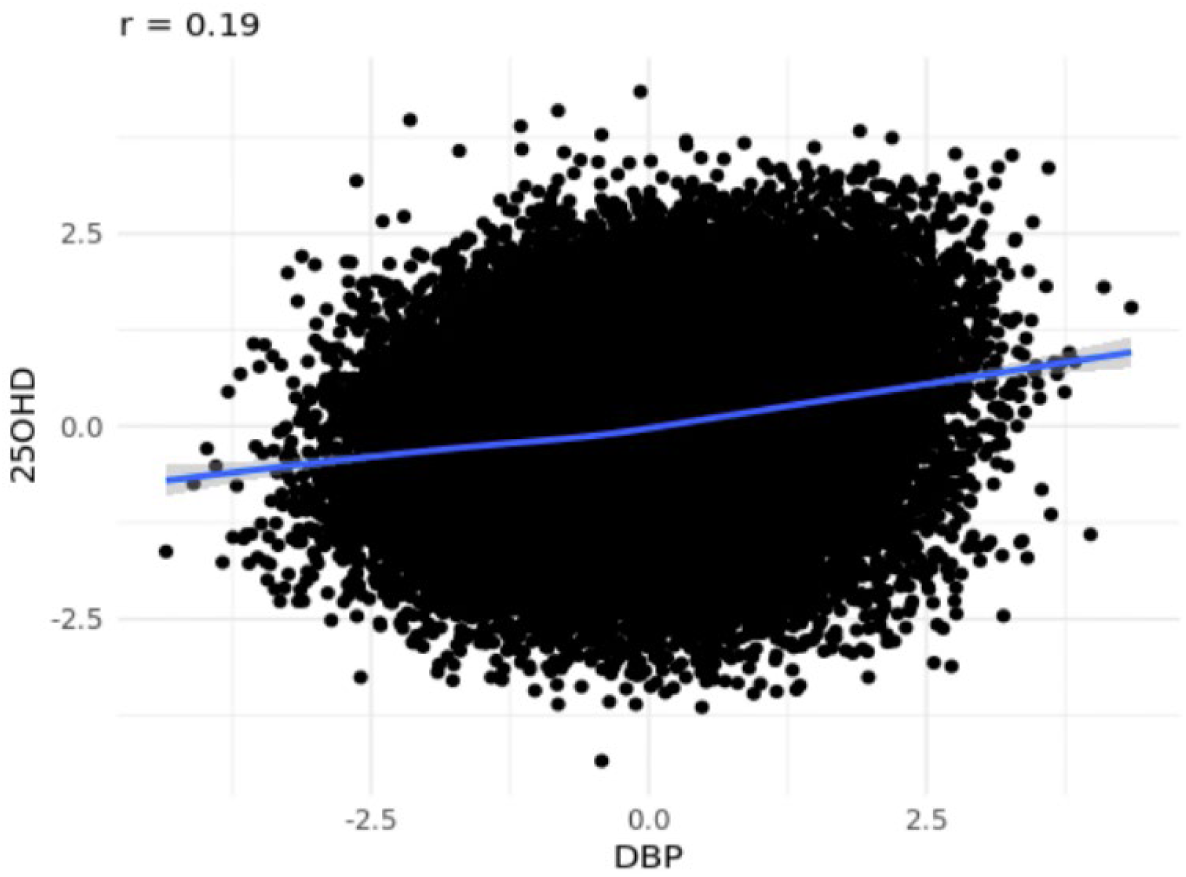
Correlation between vitamin D binding protein (DBP and 25 hydroxyvitamin D (25OHD) Scatter plot of DBP and 25OHD levels in the full sample (Pearson’s correlation coefficient r = 0.19, pvalue < 2.2e-16).

**Figure S8.**
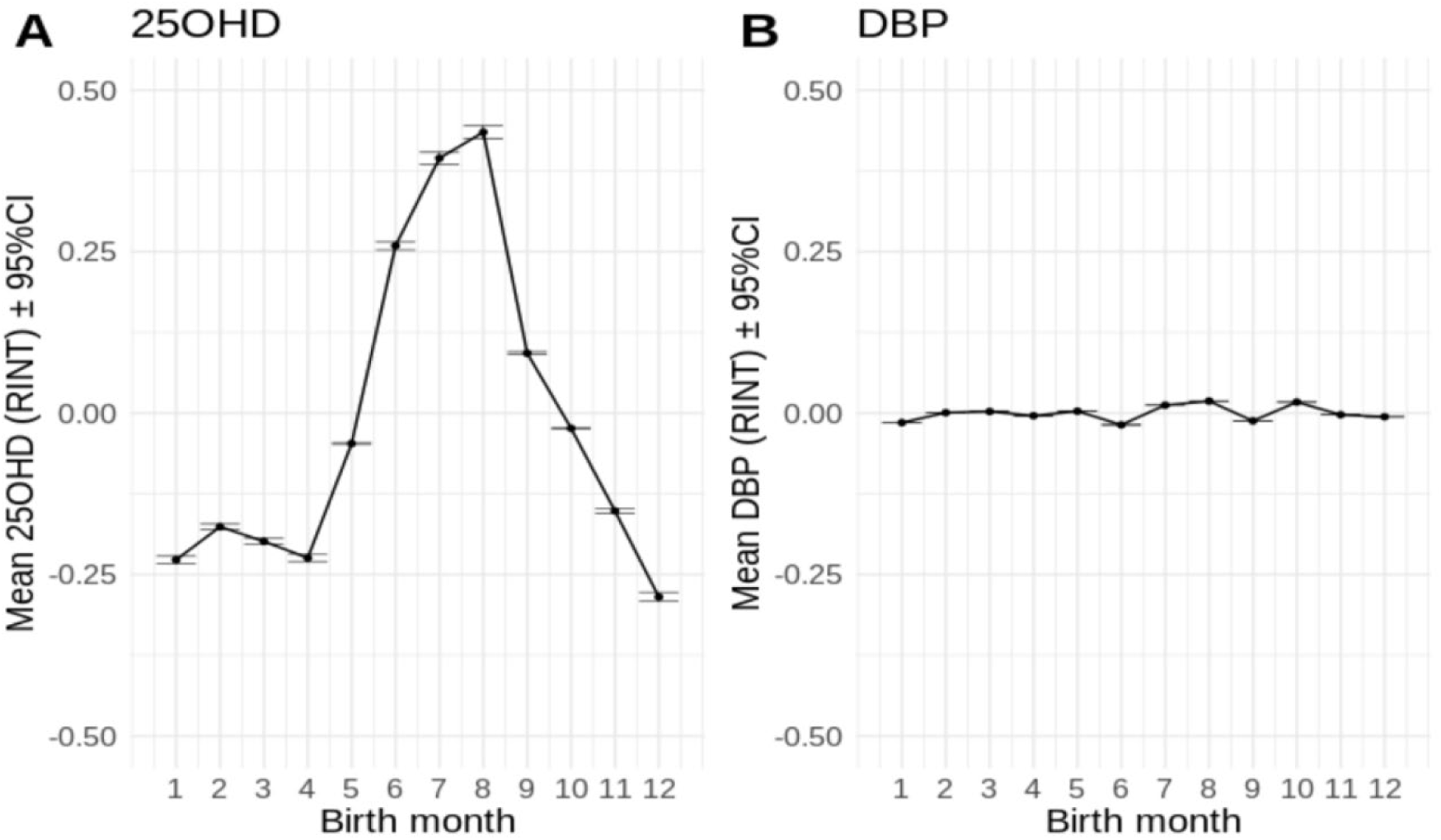
Seasonal variation – 25 hydroxyvitamin D (25OHD and vitamin D binding protein (DBP) concentration by month of birth Meant monthly concentration of 25 hydroxyvitamin D (25OHD) (panel a) and vitamin D binding protein (DBP) (panel b) by month of birth. The dried blood spots were most collected soon after birth Neonates or within 2 days of birth.

**Figure S9.**
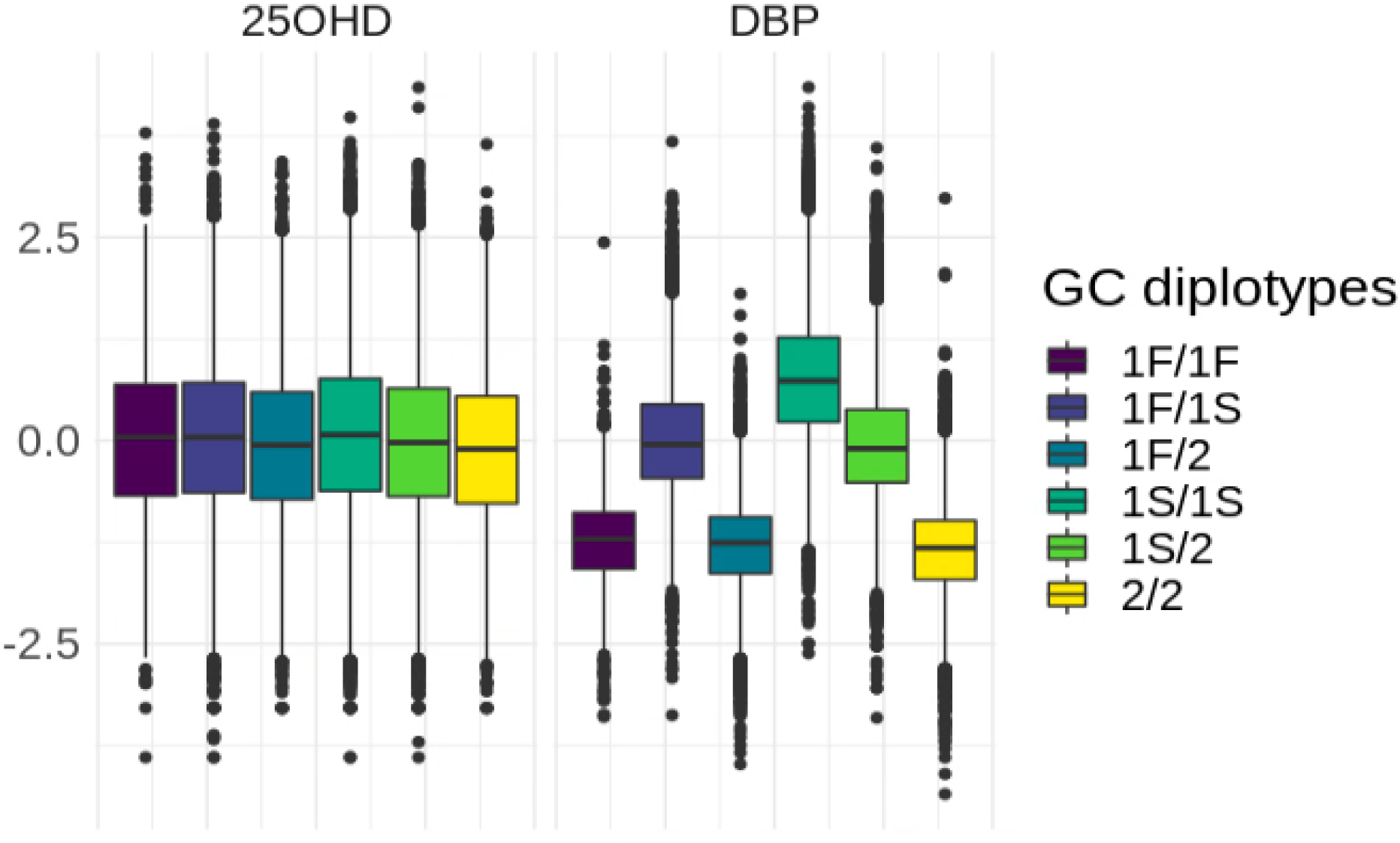
DBP and 25OHD concentration by GC haplotypes combinations (diplotypes) in the full sample (European and non-European ancestry) Distribution of 25 hydroxyvitamin D (25OHD) and vitamin D binding protein (DBP) by the six GC diplotypes in the full sample (European and non-European ancestry).

**Figure S10.**
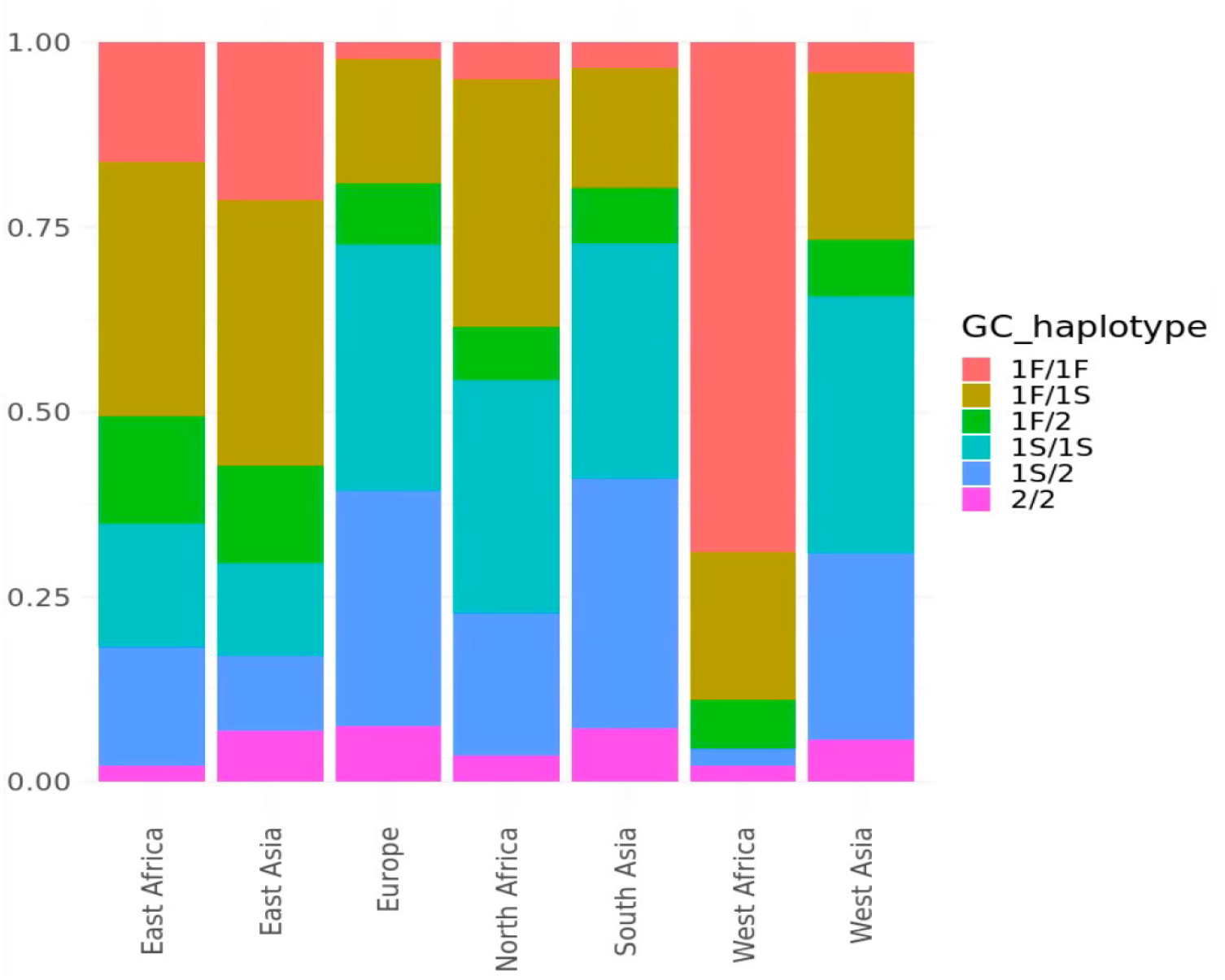
GC diplotypes by inferred ancestry groups Frequencies of GC haplotype

**Figure S11.**
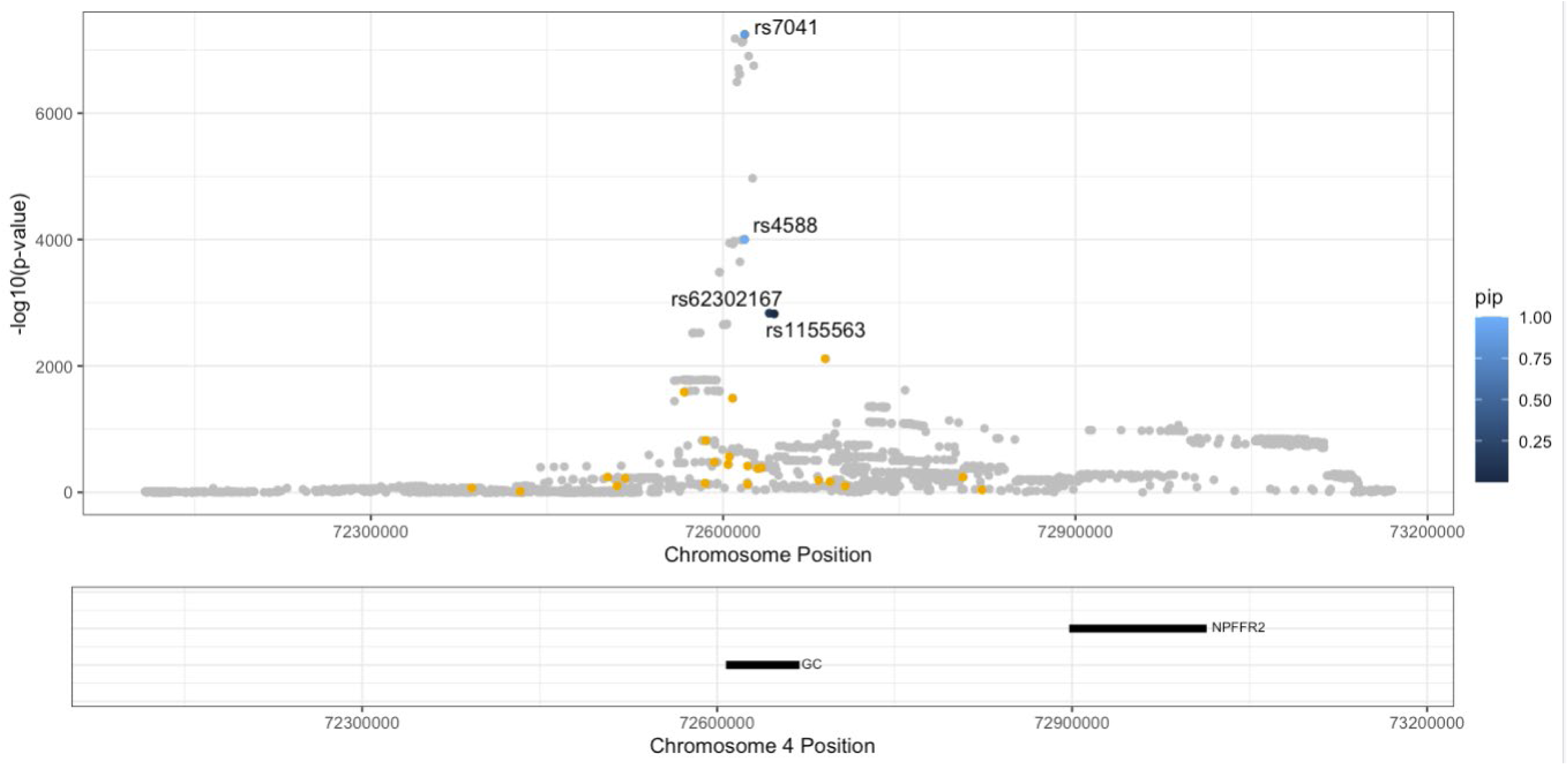
Fine mapping of candidate regions – chromosome 4 Fine-mapping results from PolyFun + SuSiE, with corresponding posterior causal probability (PIP) for a 1Mb window around the GC gene, on chromosome 4.

**Figure S12.**
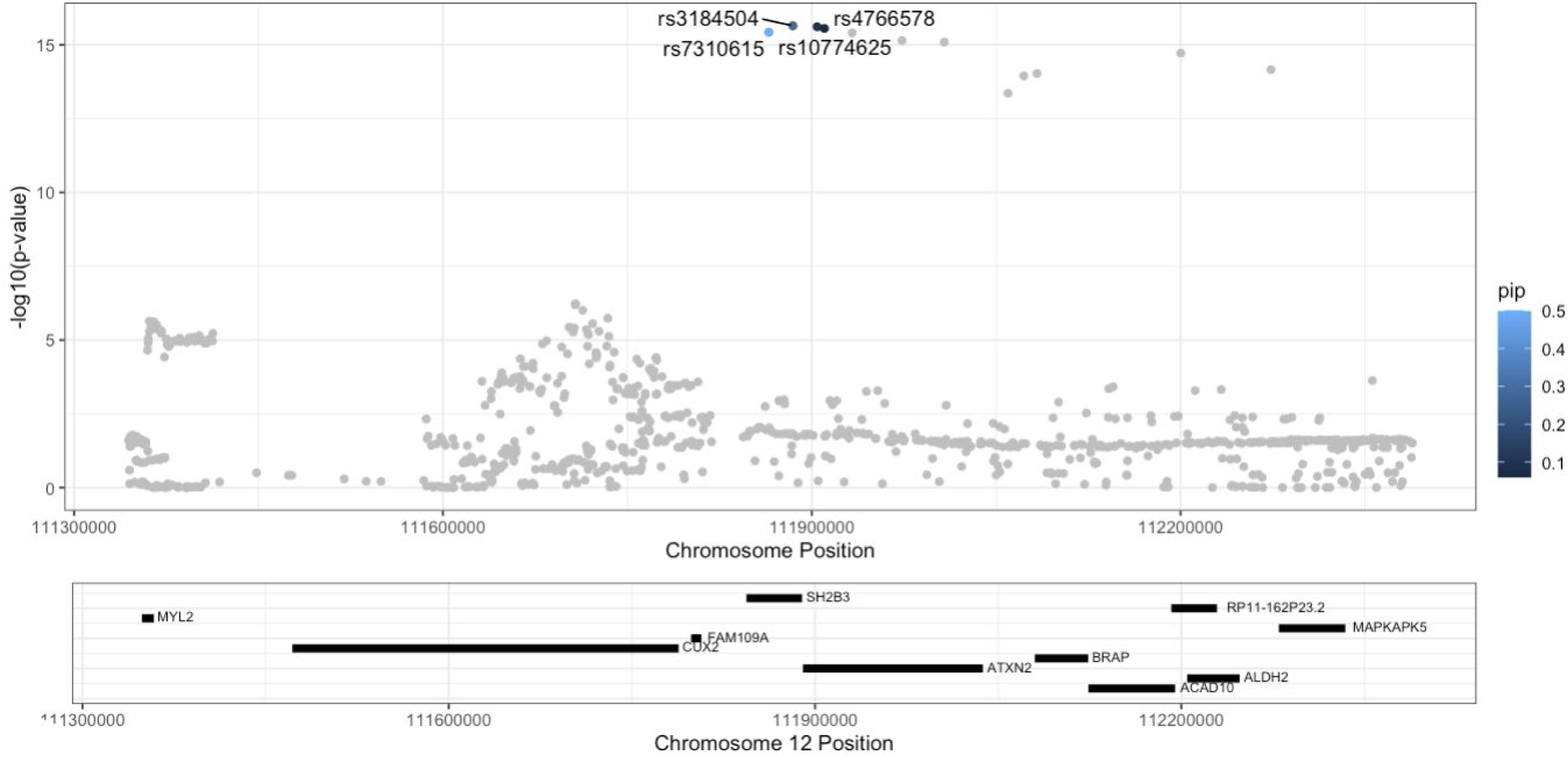
Fine-mapping of candidate region – chromosome 12 Fine-mapping results from PolyFun + SuSiE, with corresponding posterior causal probability (PIP) for a 1Mb window around the SH2B3 gene on chromosome 12.

**Figure S13.**
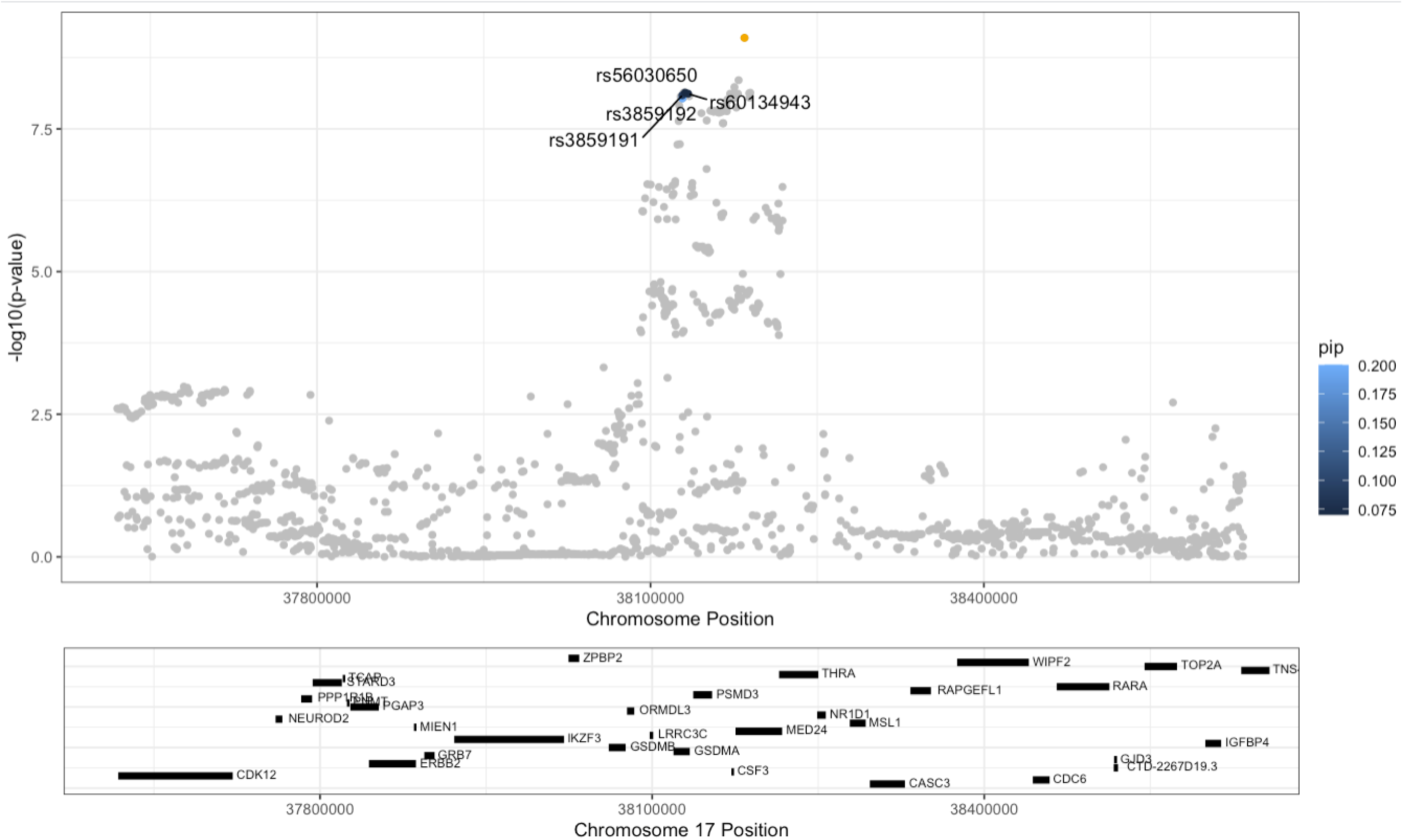
Fine-mapping of candidate region – chromosome 17 Fine-mapping results from PolyFun + SuSiE, with corresponding posterior causal probability (PIP) for a 1Mb window around the PSMD3 gene on chromosome 17.

**Figure S14.**
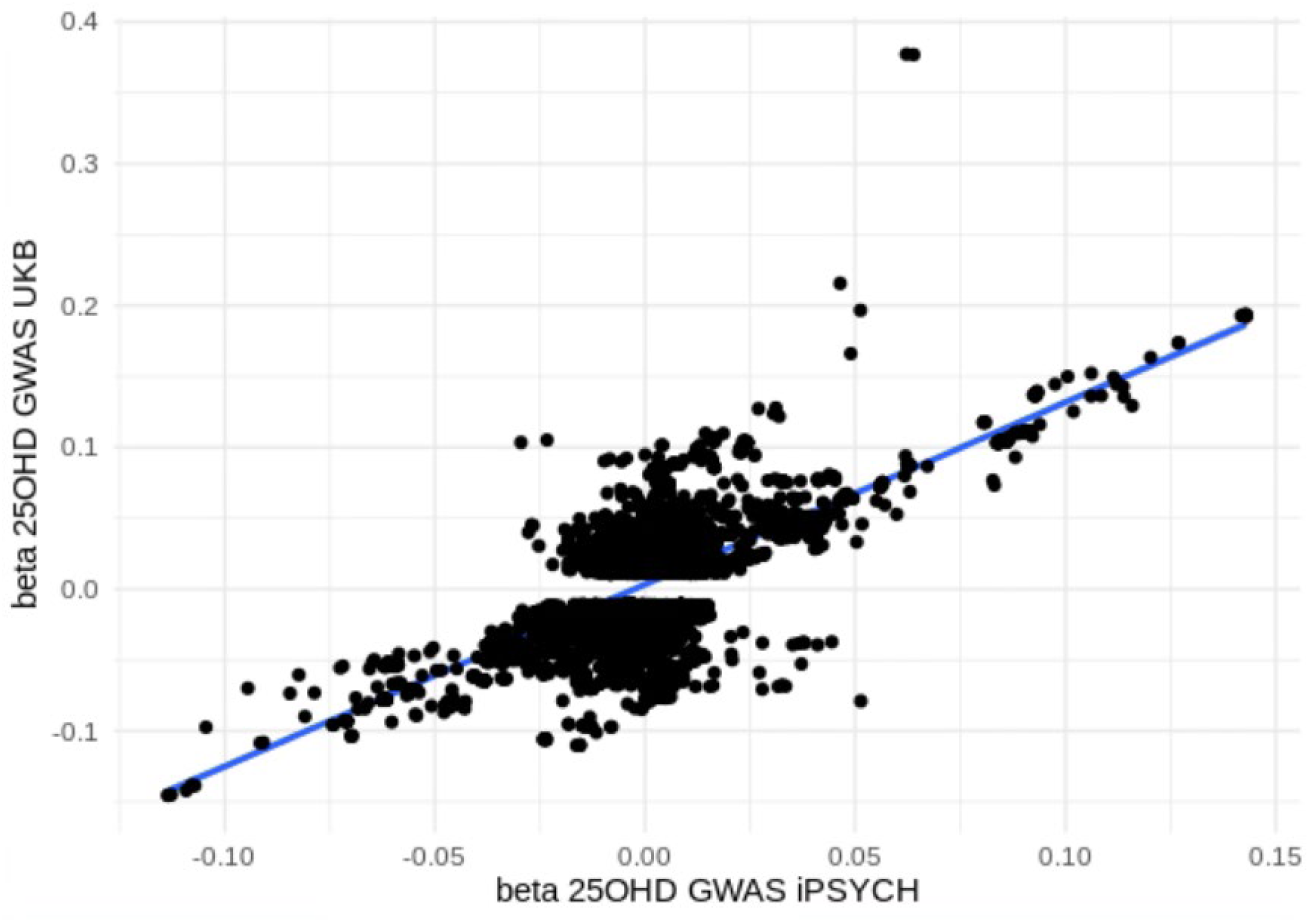
Correlation between the SNP estimates for the 25 hydroxyvitamin D (25OHD) GWAS estimates based on the iPSYCH case-cohort versus UK Biobank samples Union of the genome-wide significant SNPs (p-value < 5e-8) for the 25OHD GWAS on iPSYCH (x-axis) vs. UKB 25OHD GWAS (y-axis).

**Figure S15.**
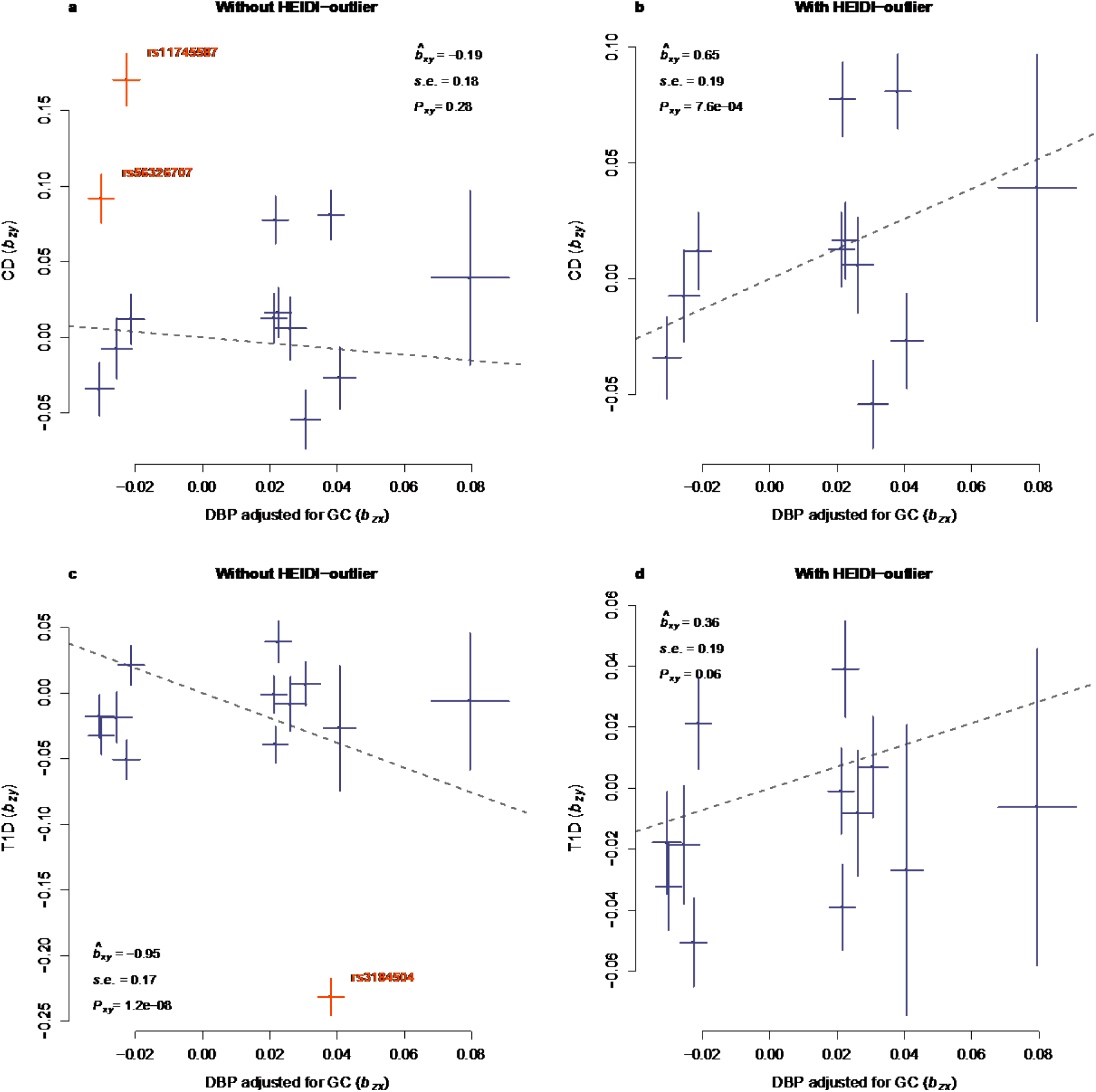
Mendelian randomization between GWAS summary statistics for DBP condition on GC (DBP_GC) versus Crohn’s disease (CD) and type 1 diabetes (T1D) GSMR analysis to test for the relationship between DBP conditional on GC and Crohn’s disease (CD) and type 1 diabetes (T1D). The GSMR estimates (dashed lines) are accompanied by estimate of effect (beta), standard error and p-value. The dash line represents GSMR estimate of effect. The top two panels show the effects of DBP_GC on CD, a) without HEIDI-outlier and b) with HEIDI-outlier respectively. We labelled and highlighted the pleiotropic SNPs identified by HEIDI- outlier in orange. Shown in the bottom row are the effects of DBP on T1D, c) without HEIDI-outlier and d) with HEIDI-outlier. The pleiotropic SNP were labelled and highlighted. The bar shown in the graph represents the GWAS standard errors at each SNP.

**Figure S16.**
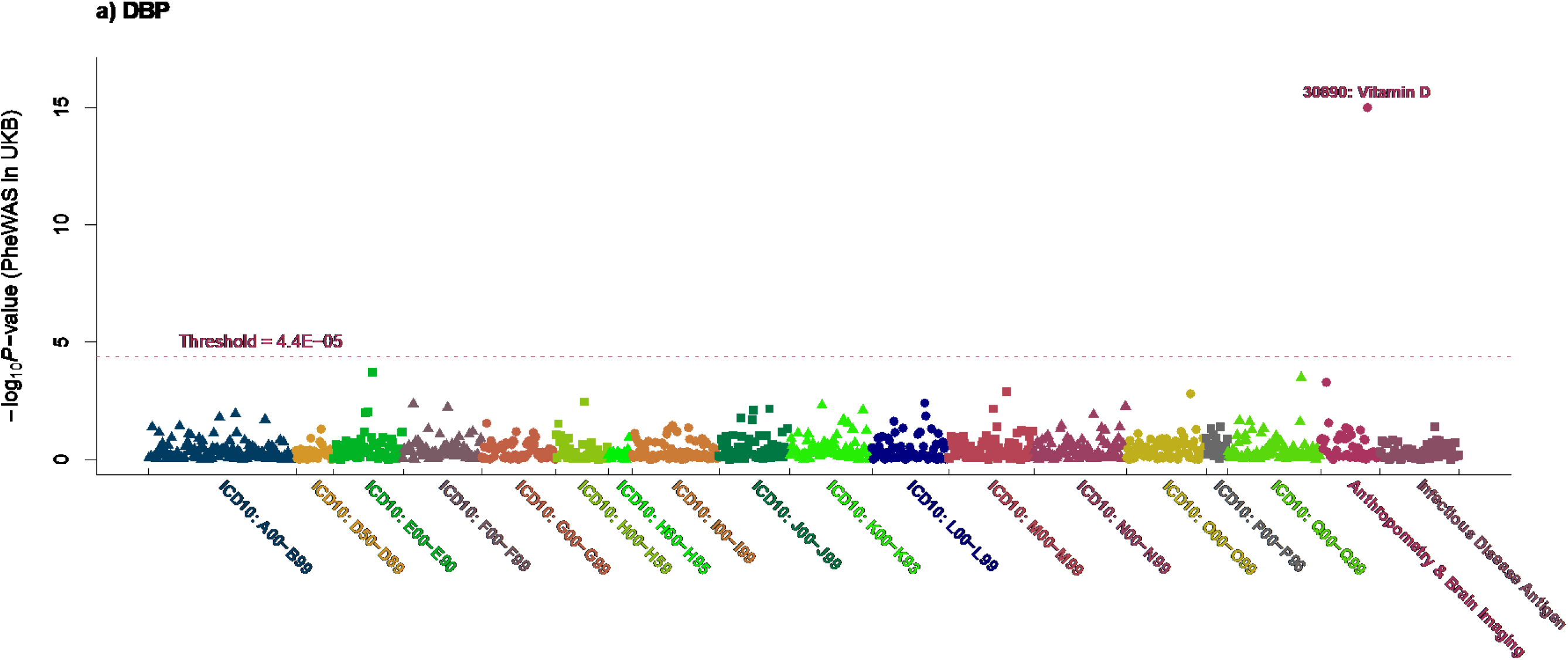
Phenome-wide association study (PheWAS) analysis of vitamin D binding protein (DBP) summary statistics in the UK Biobank PheWAS analysis of DBP concentration in UKB. Shown in the plots are the PheWAS results of DBP. The PheWAS analysis was conducted in UKB with 1 149 phenotypes, 1 027 diseases classified by ICD-10 code, 52 anthropometric and brain imaging measures, 70 infectious disease antigens. The threshold was 4.4×10^-5^. The significant phenotypes were labelled.

**Figure S17.**
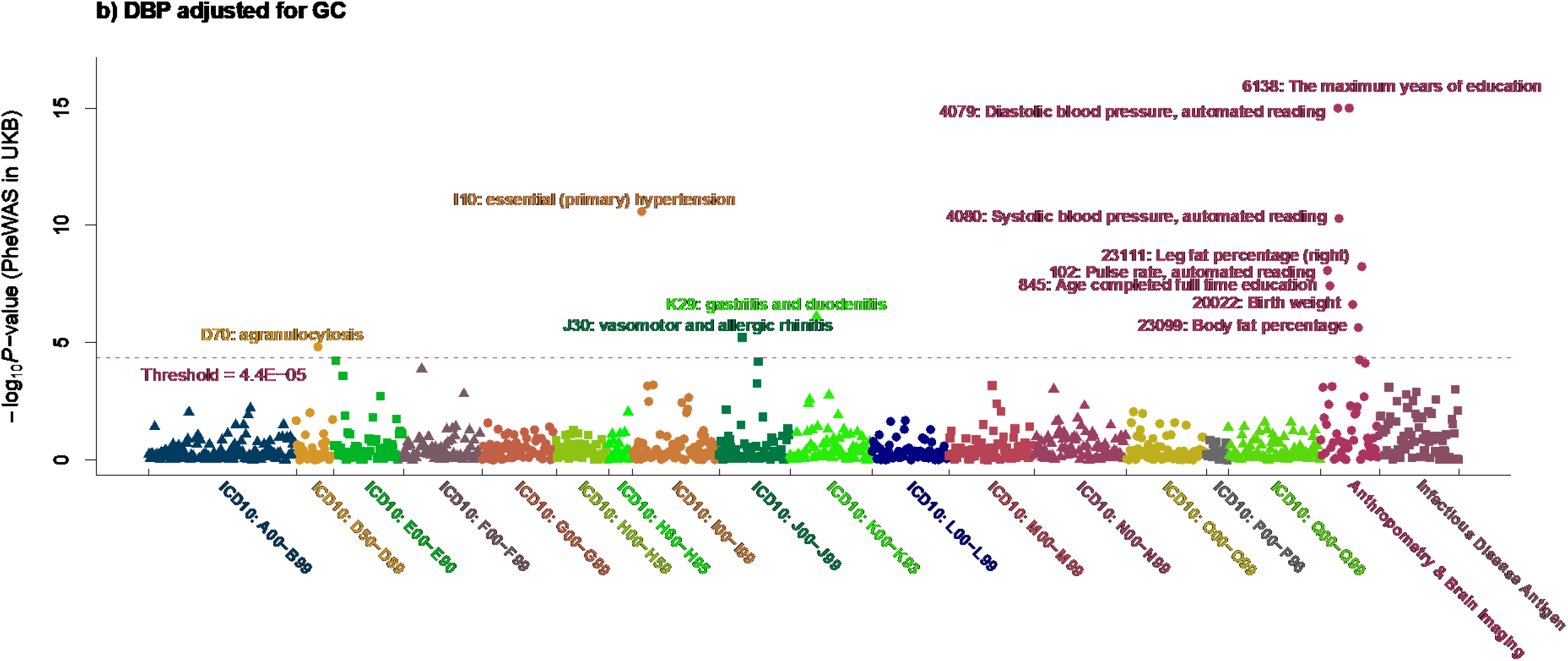
Phenome-wide association study (PheWAS) analysis of vitamin D binding protein (DBP) adjusted for GC haplotypes summary statistics in the UK Biobank PheWAS analysis of DBP concentration in UKB. Shown in the plots are the PheWAS results of DBP adjusted for GC genotypes. The PheWAS analysis was conducted in UKB with 1 149 phenotypes, 1 027 diseases classified by ICD-10 code, 52 anthropometric and brain imaging measures, 70 infectious disease antigens. The threshold was 4.4×10^-5^. The significant phenotypes were labelled.

### SUPPLEMENTARY DATA *(see Excel workbook tabs)*

SD1 Supplementary Data 1. Covariate associations.

SD2 Supplementary Data 2: GWAS SNP-based heritability estimates for 25 hydroxyvitamin D (25OHD), and vitamin D binding protein (DBP) with and without adjustment for GC haplotypes.

SD3 Supplementary Data 3. Summary of genome-wide associations estimates for 25 hydroxyvitamin D (25OHD), vitamin D binding protein (DBP) and DBP adjusted for GC (DBP_GC).

SD4 Supplementary Data 4. 143 independent associations identified with GCTA- COJO (conditional and joint) on the UKB GWAS and after conditioning on DBP, compared to the iPSYCH 25OHD GWAS

SD5 Supplementary Data 5. Supplementary Data 5. Out-of-sample variance explained by the different polygenic scores (prs), adjusted for sex, age and first 20 principal components (PCs).

SD6 Supplementary Data 6. Summary of genome-wide associations in the sub-cohort sample only

SD7 Supplementary Data 7. FUMA gene-based analysis results SD8 Supplementary Data 8. Results from FUMA gene set analysis

SD9 Supplementary Data 9: SMR results of DBP protein concentration

SD10 Supplementary Data 10: SMR results of DBP protein concentration conditional on the GC genotypes

SD11 Supplementary Data 11 SuSiE fine-mapping results.

SD12 Supplementary Data 12. Bi-directional GSMR associations between vitamin D and DBP

SD13 Supplementary Data 13 - GSMR results of DBP versus selected phenotypes SD14 Supplementary Data 14: GSMR results of DBP adjusted for the GC genotypes versus selected phenotypes

SD15 Supplementary Data 15: PheWAS analysis of the DBP concentration in UKB SD16 Supplementary Data 16: Odds ratio of DBP PRS on vitamin D deficiency

SD17 Supplementary Data 16: PheWAS analysis of the DBP_GC concentration in UKB

## References

1. Bouillon, R., Schuit, F., Antonio, L. & Rastinejad, F. Vitamin D Binding Protein: A Historic Overview. Front. Endocrinol. (Lausanne) 10, 910 (2019).

2. Chun, R.F. New perspectives on the vitamin D binding protein. Cell Biochem. Funct. 30, 445–456 (2012).

3. Mendel, C.M. The free hormone hypothesis: a physiologically based mathematical model. Endocr. Rev. 10, 232–274 (1989).

4. Bikle, D.D. & Schwartz, J. Vitamin D Binding Protein, Total and Free Vitamin D Levels in Different Physiological and Pathophysiological Conditions. Front. Endocrinol. (Lausanne) 10, 317 (2019).

5. Henderson, C.M., et al. Vitamin D-Binding Protein Deficiency and Homozygous Deletion of the GC Gene. N. Engl. J. Med. 380, 1150–1157 (2019).

6. Zella, L.A., Shevde, N.K., Hollis, B.W., Cooke, N.E. & Pike, J.W. Vitamin D-binding protein influences total circulating levels of 1,25-dihydroxyvitamin D3 but does not directly modulate the bioactive levels of the hormone in vivo. Endocrinology 149, 3656–3667 (2008).

7. Berg, A.H., et al. Development and analytical validation of a novel bioavailable 25- hydroxyvitamin D assay. PLoS One 16, e0254158 (2021).

8. Denburg, M.R., et al. Comparison of Two ELISA Methods and Mass Spectrometry for Measurement of Vitamin D-Binding Protein: Implications for the Assessment of Bioavailable Vitamin D Concentrations Across Genotypes. J. Bone Miner. Res. 31, 1128–1136 (2016).

9. Moy, K.A., et al. Genome-wide association study of circulating vitamin D-binding protein. Am. J. Clin. Nutr. 99, 1424–1431 (2014).

10. Ahn, J., et al. Genome-wide association study of circulating vitamin D levels. Hum. Mol. Genet. (2010).

11. Revez, J.A., et al. Genome-wide association study identifies 143 loci associated with 25 hydroxyvitamin D concentration. Nat Commun 11, 1647 (2020).

12. Wang, T.J., et al. Common genetic determinants of vitamin D insufficiency: a genome-wide association study. Lancet 376, 180–188 (2010).

13. Manousaki, D., et al. Genome-wide Association Study for Vitamin D Levels Reveals 69 Independent Loci. Am. J. Hum. Genet. 106, 327–337 (2020).

14. Cui, X., McGrath, J.J., Burne, T.H.J. & Eyles, D.W. Vitamin D and schizophrenia: 20 years on. Mol. Psychiatry (2021).

15. Cui, X.Y., Gooch, H., Petty, A., McGrath, J.J. & Eyles, D. Vitamin D and the brain: Genomic and non-genomic actions. Mol. Cell. Endocrinol. 453, 131–143 (2017).

16. Hahn, J., et al. Vitamin D and marine omega 3 fatty acid supplementation and incident autoimmune disease: VITAL randomized controlled trial. BMJ 376, e066452 (2022).

17. Jiang, X., Ge, T. & Chen, C.Y. The causal role of circulating vitamin D concentrations in human complex traits and diseases: a large-scale Mendelian randomization study. Sci. Rep. 11, 184 (2021).

18. Fletcher, J., et al. Autoimmune disease and interconnections with vitamin D. Endocr Connect 11 (2022).

19. Pedersen, C.B., et al. The iPSYCH2012 case-cohort sample: new directions for unravelling genetic and environmental architectures of severe mental disorders. Mol. Psychiatry 23, 6–14 (2018).

20. Pendergrass, S.A., et al. The use of phenome-wide association studies (PheWAS) for exploration of novel genotype-phenotype relationships and pleiotropy discovery. Genet. Epidemiol. 35, 410–422 (2011).

21. Thornton, L.M., et al. The Anorexia Nervosa Genetics Initiative (ANGI): Overview and methods. Contemp. Clin. Trials 74, 61–69 (2018).

22. Norgaard-Pedersen, B. & Hougaard, D.M. Storage policies and use of the Danish Newborn Screening Biobank. J. Inherit. Metab. Dis. 30, 530–536 (2007).

23. Hollegaard, M.V., et al. Whole genome amplification and genetic analysis after extraction of proteins from dried blood spots. Clin. Chem. 53, 1161–1162 (2007).

24. Boelt, S.G., et al. Sensitive and Robust LC-MS/MS Assay to Quantify 25- Hydroxyvitamin D in Leftover Protein Extract from Dried Blood Spots. International Journal of Neonatal Screening 7, 82 (2021).

25. Boelt, S.G., et al. A method to correct for the influence of bovine serum albumin- associated vitamin D metabolites in protein extracts from neonatal dried blood spots. BMC Res. Notes 15, 194 (2022).

26. Eyles, D., et al. A sensitive LC/MS/MS assay of 25OH vitamin D3 and 25OH vitamin D2 in dried blood spots. Clin. Chim. Acta 403, 145–151 (2009).

27. Kvaskoff, D., et al. Minimizing Matrix Effects for the Accurate Quantification of 25- Hydroxyvitamin D Metabolites in Dried Blood Spots by LC-MS/MS. Clin. Chem. 62, 639–646 (2016).

28. Kvaskoff, D., Ko, P., Simila, H.A. & Eyles, D.W. Distribution of 25-hydroxyvitamin D3 in dried blood spots and implications for its quantitation by tandem mass spectrometry. J. Chromatogr. B Analyt. Technol. Biomed. Life Sci. 901, 47–52 (2012).

29. Carter, G.D., et al. Hydroxyvitamin D assays: An historical perspective from DEQAS. J. Steroid Biochem. Mol. Biol. 177, 30–35 (2018).

30. McCarthy, S., et al. A reference panel of 64,976 haplotypes for genotype imputation. Nat. Genet. 48, 1279–1283 (2016).

31. Lam, M., et al. RICOPILI: Rapid Imputation for COnsortias PIpeLIne. Bioinformatics 36, 930–933 (2020).

32. Prive, F., Luu, K., Blum, M.G.B., McGrath, J.J. & Vilhjalmsson, B.J. Efficient toolkit implementing best practices for principal component analysis of population genetic data. Bioinformatics 36, 4449–4457 (2020).

33. Prive, F., et al. Portability of 245 polygenic scores when derived from the UK Biobank and applied to 9 ancestry groups from the same cohort. Am. J. Hum. Genet. 109, 12–23 (2022).

34. Yang, J., Lee, S.H., Goddard, M.E. & Visscher, P.M. GCTA: a tool for genome-wide complex trait analysis. Am. J. Hum. Genet. 88, 76–82 (2011).

35. Jiang, L., et al. A resource-efficient tool for mixed model association analysis of large- scale data. Nat. Genet. 51, 1749–1755 (2019).

36. Yang, J., et al. Conditional and joint multiple-SNP analysis of GWAS summary statistics identifies additional variants influencing complex traits. Nat. Genet. 44, 369–375, S361-363 (2012).

37. Zhu, Z., et al. Causal associations between risk factors and common diseases inferred from GWAS summary data. Nat Commun 9, 224 (2018).

38. Yengo, L., et al. Meta-analysis of genome-wide association studies for height and body mass index in approximately 700000 individuals of European ancestry. Hum. Mol. Genet. 27, 3641–3649 (2018).

39. Zaitlen, N., et al. Using extended genealogy to estimate components of heritability for 23 quantitative and dichotomous traits. PLoS Genet. 9, e1003520 (2013).

40. Bulik-Sullivan, B.K., et al. LD Score regression distinguishes confounding from polygenicity in genome-wide association studies. Nat. Genet. 47, 291–295 (2015).

41. Zeng, J., et al. Widespread signatures of natural selection across human complex traits and functional genomic categories. Nat Commun 12, 1164 (2021).

42. Privé, F., Arbel, J. & Vilhjálmsson, B.J. LDpred2: better, faster, stronger. Bioinformatics 36, 5424–5431 (2020).

43. Weissbrod, O., et al. Functionally informed fine-mapping and polygenic localization of complex trait heritability. Nat. Genet. 52, 1355–1363 (2020).

44. Zou, Y., Carbonetto, P., Wang, G. & Stephens, M. Fine-mapping from summary data with the “Sum of Single Effects” model. bioRxiv, 2021.2011.2003.467167 (2022).

45. Gazal, S., et al. Linkage disequilibrium-dependent architecture of human complex traits shows action of negative selection. Nat. Genet. 49, 1421–1427 (2017).

46. McLaren, W., et al. The Ensembl Variant Effect Predictor. Genome Biol. 17, 122 (2016).

47. Lloyd-Jones, L.R., et al. Improved polygenic prediction by Bayesian multiple regression on summary statistics. Nat Commun 10, 5086 (2019).

48. Lambert, S.A., et al. The Polygenic Score Catalog as an open database for reproducibility and systematic evaluation. Nat. Genet. 53, 420–425 (2021).

49. Bulik-Sullivan, B., et al. An atlas of genetic correlations across human diseases and traits. Nat. Genet. 47, 1236–1241 (2015).

50. Watanabe, K., Taskesen, E., van Bochoven, A. & Posthuma, D. Functional mapping and annotation of genetic associations with FUMA. Nat Commun 8, 1826 (2017).

51. GTEx Consortium. Genetic effects on gene expression across human tissues. Nature 550, 204 (2017).

52. Institute of Medicine. Dietary Reference Intakes for Calcium and Vitamin D, (National Academies Press, 2010).

53. Ashley, B., et al. Placental uptake and metabolism of 25(OH)vitamin D determine its activity within the fetoplacental unit. Elife 11 (2022).

54. Auburger, G., et al. 12q24 locus association with type 1 diabetes: SH2B3 or ATXN2? World J. Diabetes 5, 316–327 (2014).

55. Liu, X., Xia, S., Zhang, Z., Wu, H. & Lieberman, J. Channelling inflammation: gasdermins in physiology and disease. Nature Reviews Drug Discovery 20, 384–405 (2021).

56. Bjorkhem-Bergman, L., Torefalk, E., Ekstrom, L. & Bergman, P. Vitamin D binding protein is not affected by high-dose vitamin D supplementation: a post hoc analysis of a randomised, placebo-controlled study. BMC Res. Notes 11, 619 (2018).

57. Chun, R.F., et al. Vitamin D and DBP: the free hormone hypothesis revisited. J. Steroid Biochem. Mol. Biol. 144 Pt A, 132-137 (2014).

58. Banerjee, R.R., et al. Very Low Vitamin D in a Patient With a Novel Pathogenic Variant in the GC Gene That Encodes Vitamin D-Binding Protein. *J Endocr Soc* 5, bvab104 (2021).

59. Jones, K.S., et al. 25(OH)D2 half-life is shorter than 25(OH)D3 half-life and is influenced by DBP concentration and genotype. J. Clin. Endocrinol. Metab. 99, 3373–3381 (2014).

60. Harroud, A. & Richards, J.B. Mendelian randomization in multiple sclerosis: A causal role for vitamin D and obesity? Mult. Scler. 24, 80–85 (2018).

61. Manousaki, D., et al. Low-Frequency Synonymous Coding Variation in CYP2R1 Has Large Effects on Vitamin D Levels and Risk of Multiple Sclerosis. Am. J. Hum. Genet. 101, 227–238 (2017).

62. Nielsen, N.M., et al. Neonatal vitamin D status and risk of multiple sclerosis: A population-based case-control study. Neurology 88, 44–51 (2017).

63. Rhead, B., et al. Mendelian randomization shows a causal effect of low vitamin D on multiple sclerosis risk. Neurol Genet 2, e97 (2016).

64. Mokry, L.E., et al. Vitamin D and Risk of Multiple Sclerosis: A Mendelian Randomization Study. PLoS Med. 12, e1001866 (2015).

65. Deluca, G.C., Kimball, S.M., Kolasinski, J., Ramagopalan, S.V. & Ebers, G.C. The Role of Vitamin D in Nervous System Health and Disease. Neuropathol. Appl. Neurobiol. (2013).

66. Cutolo, M., Otsa, K., Uprus, M., Paolino, S. & Seriolo, B. Vitamin D in rheumatoid arthritis. Autoimmunity Reviews 7, 59–64 (2007).

67. Merlino, L.A., et al. Vitamin D intake is inversely associated with rheumatoid arthritis: results from the Iowa Women’s Health Study. Arthritis Rheum. 50, 72–77 (2004).

68. Hewison, M. Vitamin D and the immune system. J. Endocrinol. 132, 173–175 (1992).

69. Xie, Z., Wang, X. & Bikle, D.D. Editorial: Vitamin D Binding Protein, Total and Free Vitamin D Levels in Different Physiological and Pathophysiological Conditions. Front. Endocrinol. (Lausanne) 11, 40 (2020).

70. Jassil, N.K., Sharma, A., Bikle, D. & Wang, X. Vitamin D Binding Protein and 25- Hydroxyvitamin D Levels: Emerging Clinical Applications. Endocr. Pract. 23, 605–613 (2017).

71. Nielson, C.M., et al. Free 25-Hydroxyvitamin D: Impact of Vitamin D Binding Protein Assays on Racial-Genotypic Associations. J. Clin. Endocrinol. Metab. 101, 2226–2234 (2016).

72. Powe, C.E., et al. Vitamin D-binding protein and vitamin D status of black Americans and white Americans. N. Engl. J. Med. 369, 1991–2000 (2013).

73. Vitamin D–Binding Protein and Vitamin D in Blacks and Whites. N. Engl. J. Med. 370, 878–881 (2014).

74. Alzaman, N.S., Dawson-Hughes, B., Nelson, J., D’Alessio, D. & Pittas, A.G. Vitamin D status of black and white Americans and changes in vitamin D metabolites after varied doses of vitamin D supplementation. Am. J. Clin. Nutr. 104, 205–214 (2016).

75. Pietzner, M., et al. Mapping the proteo-genomic convergence of human diseases. Science 374, eabj1541 (2021).

76. Ferkingstad, E., et al. Large-scale integration of the plasma proteome with genetics and disease. Nat. Genet. 53, 1712–1721 (2021).

77. Buniello, A., et al. The NHGRI-EBI GWAS Catalog of published genome-wide association studies, targeted arrays and summary statistics 2019. Nucleic Acids Res. 47, D1005–D1012 (2019).

78. Kronenberg, F., et al. Influence of hematocrit on the measurement of lipoproteins demonstrated by the example of lipoprotein(a). Kidney Int. 54, 1385–1389 (1998).

79. Hall, E.M., Flores, S.R. & De Jesus, V.R. Influence of Hematocrit and Total-Spot Volume on Performance Characteristics of Dried Blood Spots for Newborn Screening. Int J Neonatal Screen 1, 69–78 (2015).

## SUPPLEMENTARY REFERENCES

1. Carter, G.D., et al. Hydroxyvitamin D assays: An historical perspective from DEQAS. J. Steroid Biochem. Mol. Biol. 177, 30–35 (2018).

2. Trubetskoy, V., et al. Mapping genomic loci implicates genes and synaptic biology in schizophrenia. Nature 604, 502–508 (2022).

3. Howard, D.M., et al. Genome-wide meta-analysis of depression identifies 102 independent variants and highlights the importance of the prefrontal brain regions. Nat. Neurosci. 22, 343–352 (2019).

4. Mullins, N., et al. Genome-wide association study of more than 40,000 bipolar disorder cases provides new insights into the underlying biology. Nat. Genet. 53, 817–829 (2021).

5. Grove, J., et al. Identification of common genetic risk variants for autism spectrum disorder. Nat. Genet. 51, 431–444 (2019).

6. Demontis, D., et al. Discovery of the first genome-wide significant risk loci for attention deficit/hyperactivity disorder. Nat. Genet. 51, 63–75 (2019).

7. Marioni, R.E., et al. GWAS on family history of Alzheimer’s disease. Transl Psychiatry 8, 99 (2018).

8. Okbay, A., et al. Polygenic prediction of educational attainment within and between families from genome-wide association analyses in 3 million individuals. Nat. Genet. 54, 437–449 (2022).

9. International Multiple Sclerosis Genetics, C. Multiple sclerosis genomic map implicates peripheral immune cells and microglia in susceptibility. Science 365(2019).

10. van Rheenen, W., et al. Common and rare variant association analyses in amyotrophic lateral sclerosis identify 15 risk loci with distinct genetic architectures and neuron-specific biology. Nat. Genet. 53, 1636–1648 (2021).

11. Chiou, J., et al. Interpreting type 1 diabetes risk with genetics and single-cell epigenomics. Nature 594, 398–402 (2021).

12. de Lange, K.M., et al. Genome-wide association study implicates immune activation of multiple integrin genes in inflammatory bowel disease. Nat. Genet. 49, 256–261 (2017).

13. Okada, Y., et al. Genetics of rheumatoid arthritis contributes to biology and drug discovery. Nature 506, 376–381 (2014).

